# Optimal SARS-CoV-2 vaccine allocation using real-time seroprevalence estimates in Rhode Island and Massachusetts

**DOI:** 10.1101/2021.01.12.21249694

**Authors:** Thu Nguyen-Anh Tran, Nathan Wikle, Joseph Albert, Haider Inam, Emily Strong, Karel Brinda, Scott M Leighow, Fuhan Yang, Sajid Hossain, Justin R Pritchard, Philip Chan, William P Hanage, Ephraim M Hanks, Maciej F Boni

## Abstract

As three SARS-CoV-2 vaccines come to market in Europe and North America in the winter of 2020-2021, distribution networks will be in a race against a major epidemiological wave of SARS-CoV-2 that began in autumn 2020. Rapid and optimized vaccine allocation is critical during this time. With 95% efficacy reported for two of the vaccines, near-term public health needs require that distribution is prioritized to the elderly, health-care workers, teachers, essential workers, and individuals with co-morbidities putting them at risk of severe clinical progression. Here, we evaluate various age-based vaccine distributions using a validated mathematical model based on current epidemic trends in Rhode Island and Massachusetts. We allow for varying waning efficacy of vaccine-induced immunity, as this has not yet been measured. We account for the fact that known COVID-positive cases may not be included in the first round of vaccination. And, we account for current age-specific immune patterns in both states. We find that allocating a substantial proportion (*>* 75%) of vaccine supply to individuals over the age of 70 is optimal in terms of reducing total cumulative deaths through mid-2021. As we do not explicitly model other high mortality groups, this result on vaccine allocation applies to all groups at high risk of mortality if infected. Our analysis confirms that for an easily transmissible respiratory virus, allocating a large majority of vaccinations to groups with the highest mortality risk is optimal. Our analysis assumes that health systems during winter 2020-2021 have equal staffing and capacity to previous phases of the SARS-CoV-2 epidemic; we do not consider the effects of understaffed hospitals or unvaccinated medical staff. Vaccinating only seronegative individuals avoids redundancy in vaccine use on individuals that may already be immune, and will result in 1% to 2% reductions in cumulative hospitalizations and deaths by mid-2021. Assuming high vaccination coverage (*>* 28%) and no major relaxations in distancing, masking, gathering size, or hygiene guidelines between now and spring 2021, our model predicts that a combination of vaccination and population immunity will lead to low or near-zero transmission levels by the second quarter of 2021.

## 1 Introduction

The international effort to bring a SARS-CoV-2 vaccine to market began in January 2020 with the release of the viral genome sequence [1]. Development of two mRNA vaccines and dozens of other vaccine candidates began shortly thereafter [2, 3], and two vaccine candidates (Moderna and Pfizer/BioNTech) were approved for use in the United States by the US Food and Drug Administration in December 2020 [4, 5]. Roll-out of these two vaccines in the US is the best near-term hope of stopping the epidemic by spring/summer 2021 and keeping the total death toll in the United States below half a million. The Centers for Disease Control and Prevention [6–8] together with state-level departments of health [9, 10] have developed vaccine distribution and prioritization plans for the first batch of doses and the first few months of distribution.

In vaccination campaigns, prioritization of certain population groups over others can influence the success of the campaign. The classic trade-off in vaccine distribution programs is between vaccinating high-contact versus high-risk individuals [11–14], and the optimal approach depends on (1) the outcome measure being used – e.g. case numbers, hospitalizations, or deaths, (2) the vaccine supply, (3) the mortality rate in the at-risk age groups, and (4) the current level of transmission. For long-term planning in influenza vaccination, there is empirical evidence that vaccination of children (the high-contact group) can lead to reduced case numbers, morbidity, and deaths for all age groups [15]; however, short-term planning is much more sensitive to small changes in roll-out details and the current state of the epidemic. The benefit of reduced transmission may come at a delay from the start of the vaccination campaign, and thus it is often safer to protect vulnerable groups with the direct benefit of vaccination rather than the indirect benefits of vaccinating others. This is especially true for the COVID-19 pandemic as SARS-CoV-2 infections have an infection mortality rate that is *>*10 times higher than that of influenza virus [16].

Vaccinating populations with real-time information on seroprevalence allows a public health system to introduce efficiencies into vaccine allocation. Here, we evaluate different age-distributions for vaccine roll-out given current age-stratified attack-rate estimates in Rhode Island and Massachusetts [17]. While we assume that confirmed seropositive individuals (i.e. those who were confirmed COVID-19 cases in the past) will not receive the vaccine in the first rounds of vaccine distribution, this will understandably be implemented through voluntary compliance as it will not be possible to have a ‘seronegatives only’ vaccine program in place due to the rapid schedule of shipping, deployment, staffing, and vaccination. The current seroprevalence also factors into allocation decisions as the pace of vaccination and simultaneous transmission will determine how quickly the population approaches the herd-immunity threshold, an approach that has beneficial non-linear effects in reducing the final tally of infected individuals [18–20].

Using a mathematical model whose fit to Rhode Island and Massachusetts COVID-19 data was described in Wikle et al [17], we consider seven different vaccine efficacy profiles/halflives (currently an unknown), we evaluate the individual importance of each 10-year age band to vaccination outcomes, we compare several common age allocations under assumptions of high and low vaccine supply, and we evaluate the magnitude of population-level effects if vaccination is dependent on serostatus. We assume that health care workers and front-line medical staff are vaccinated first, as our model is not able to evaluate the effects of a stressed and understaffed health system.

## 2 Methods

### 2.1 Model and Fitting

We use a mathematical model developed by Wikle et al [17] and fit to Rhode Island and Massachusetts data so that vaccination campaigns can be evaluated in the context of the current number of individuals already infected. As of Nov 30, total estimated attack rates are 20.7% (95% CI : 17.3% *−* 24.1%) in Rhode Island and 12.5% (95% CI : 11.5% *−* 13.5%) in Massachusetts [21]. We upgraded the model parameterization by using age-contact matrices measured in Belgium (CoMix) which are representative of lockdown and post-lockdown mixing patterns in western countries [22]. The eight age-bands in the CoMix data [22] were transformed into our nine age-bands using the `socialmixr` R package [23]. Using median values from posteriors from the new model fit, the new model’s Nov 30 attack rates are 16.8% for RI and 16.5% for MA.

To model vaccination with waning vaccine efficacy, we add a 24-stage vaccinated state (classes *Z*_1_ to *Z*_24_ in Figure 1) in order to to allow vaccine efficacy to be modelled as *x*% efficacy at *n* weeks post vaccination. Clearly, 24 vaccine efficacy estimates for different time points have not been published, but we use this 24-compartment chain to model the expected gradual changes in vaccine efficacy from the early and measured stages in clinical trials (60 to 90 days after vaccination, with observed efficacy around 95%) to stages 12 or 18 months later when the vaccine is assumed to have an exponentially waned efficacy (several parameterizations are explored). Even for early-stage efficacy, these data are not yet available because we do not know the average enrollment duration of patients in the two key trials. Nevertheless, we make several reasonable choices for shapes of this efficacy function based on the enrollment dates of each trial [24, 25]. The efficacy curves in Figure 2 are parameterized with a Hill-like function, HL^*s*^*/*(HL^*s*^ + *T*^*s*^), where HL is the efficacy half-life, *s* is the slope, and *T* is the time post-vaccination. To translate this individual efficacy into a population-level model, we define the vaccine efficacy (VE_*j*_) in the vaccinated class *Z*_*j*_ as

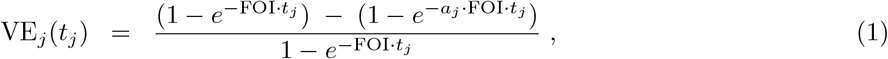

where *t*_*j*_ is the time after the final vaccine dose, FOI is the force of infection in the population, and *a*_*j*_ is the relative reduction in susceptibility for a vaccinated individual in class *Z*_*j*_ (which is *t*_*j*_ days after the final dose of vaccination). We assume an FOI of 0.001 per day, and for *t*_*j*_ = 60 or *t*_*j*_ = 90 days we assume 95% efficacy and solve for *a*_*j*_. For other values of *t*_*j*_, the vaccine efficacy is assumed to follow one of the seven patterns in Figure 2, and 24 *a*_*j*_ values (relative risk parameters for a fixed time point after vaccination) are solved for. We assume the force of infection from December 1 onward is constant and we explore three transmission scenarios. In the low-transmission scenario, transmission levels in Rhode Island and Massachusetts revert to their late spring and summer levels. Under medium transmission, transmission levels are set to their September through November mean value. Under high transmission, wintertime transmission levels continue through spring 2021; see Figure 3. The medium transmission scenario is explored in the main text, and figures for the high and low scenarios are included in the Supplementary Materials.

**Figure 1:**
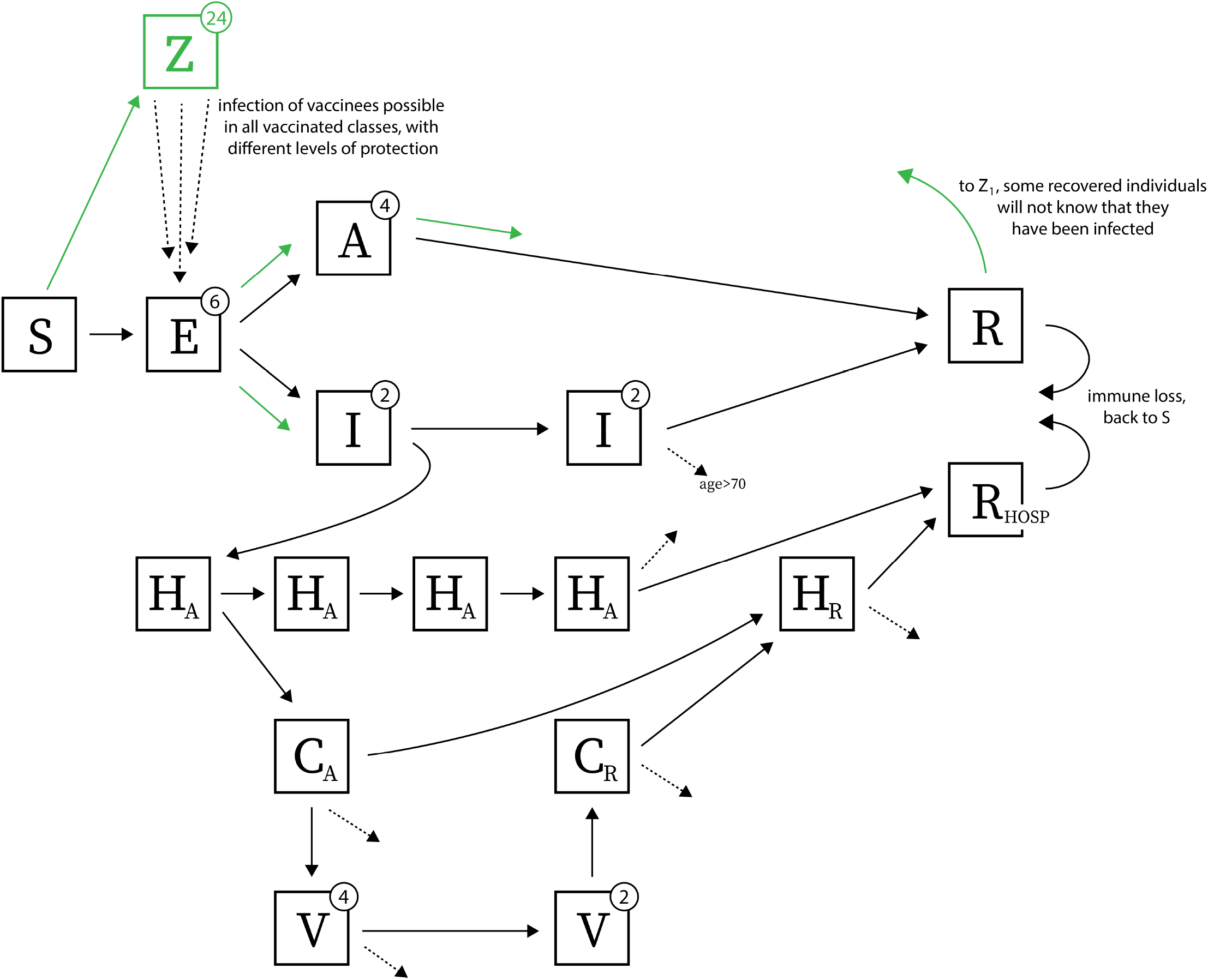
We extend the model from [17] to include a 24-stage vaccinated compartment, denoted by *Z*. Other compartments are susceptible (*S*), exposed (*E*), asymptomatic (*A*), infected and symptomatic but not hospitalized (*I*), hospitalized in acute stage of infection (*H*_*A*_), in critical care (acute stage) (*C*_*A*_), on mechanical ventilation (*V*), in critical care recovering phase (*C*_*R*_), hospitalized and recovering (*H*_*R*_), recovered (*R*), recovered from hospitalization (*R*_HOSP_). Green arrows show progression after an individual is vaccinated; dashed arrows indicate death. Exposed and asymptomatic individuals who are vaccinated receive no benefit from the vaccine and progress on their normal infection/disease course. We assume vaccines are not given to individuals with past confirmed infection. However, if no serological test is performed before vaccinating, it is possible that individuals in exposed (*E*), asymptomatic (*A*), and recovered (*R*) classes will have an equal chance of receiving the vaccine as truly susceptible individuals. Re-infection is possible as shown by arrows going from recovered *R* and *R*_HOSP_ back to susceptible compartment *S*. Vaccinees in the *Z* class are not fully protected from infection since we allow waning of vaccine-induced immunity over time; infection of vaccinated individuals is shows with the dashed arrows from *Z* to *E*.

**Figure 2:**
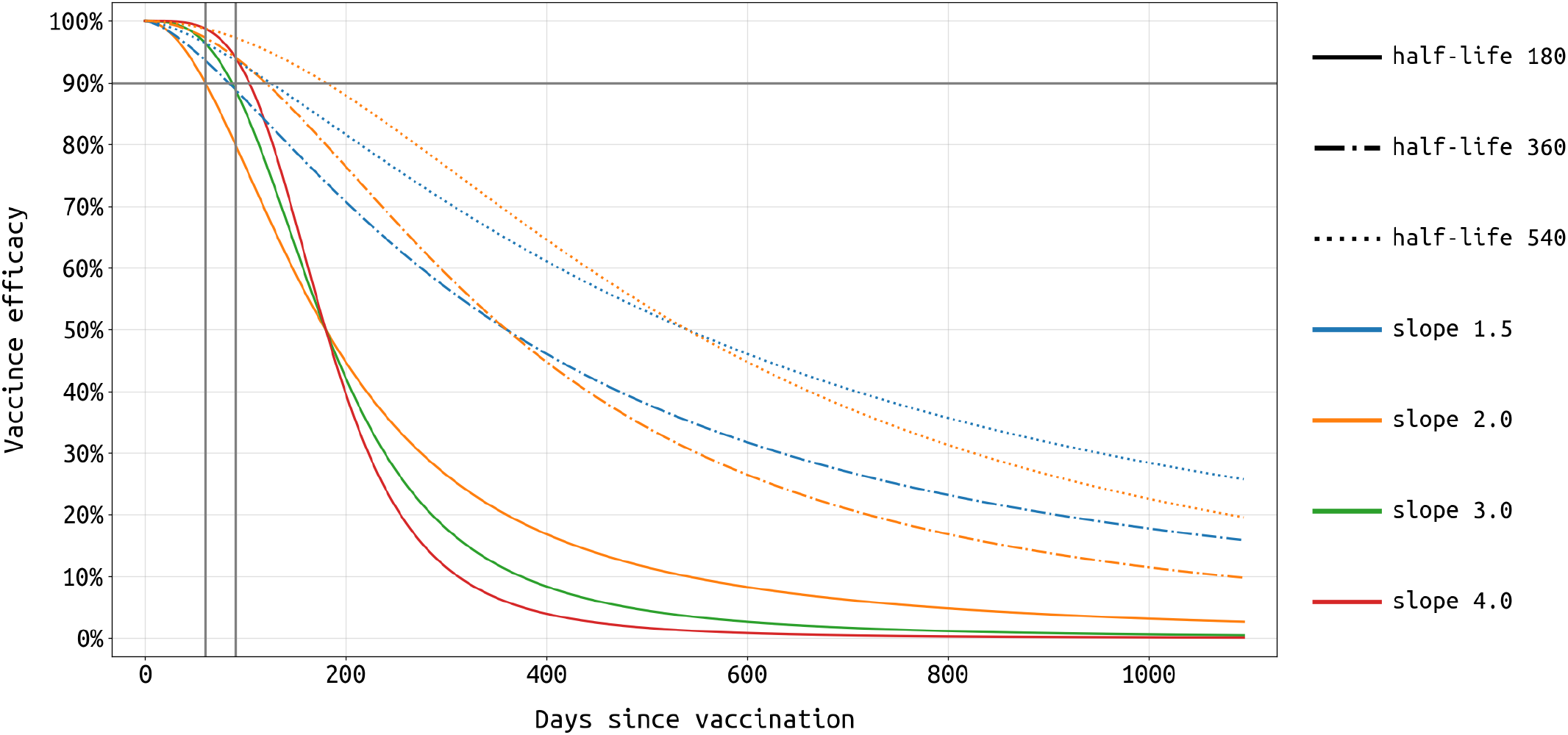
We assume that vaccine efficacy wanes over time following a Hill-like function characterized by an efficacy half-life and slope (Equation 1). The half-life (as shown with different line styles) denotes on which day after vaccination the efficacy drops below the 50% mark. The slope (shown with different line colors) denotes how fast the vaccine loses its efficacy; for example, considering the solid lines in this figure (half-life = 180), efficacy of the orange vaccine (slope = 2) reduces from 70% to 50% in 64 days while that of the red vaccine (slope = 4) does so in 36 days. Here, we choose 7 vaccine profiles whose 60-day and/or 90-day efficacies are at least 90% to represent the currently approved vaccines from Pfizer/BioNTech and Moderna.

**Figure 3:**
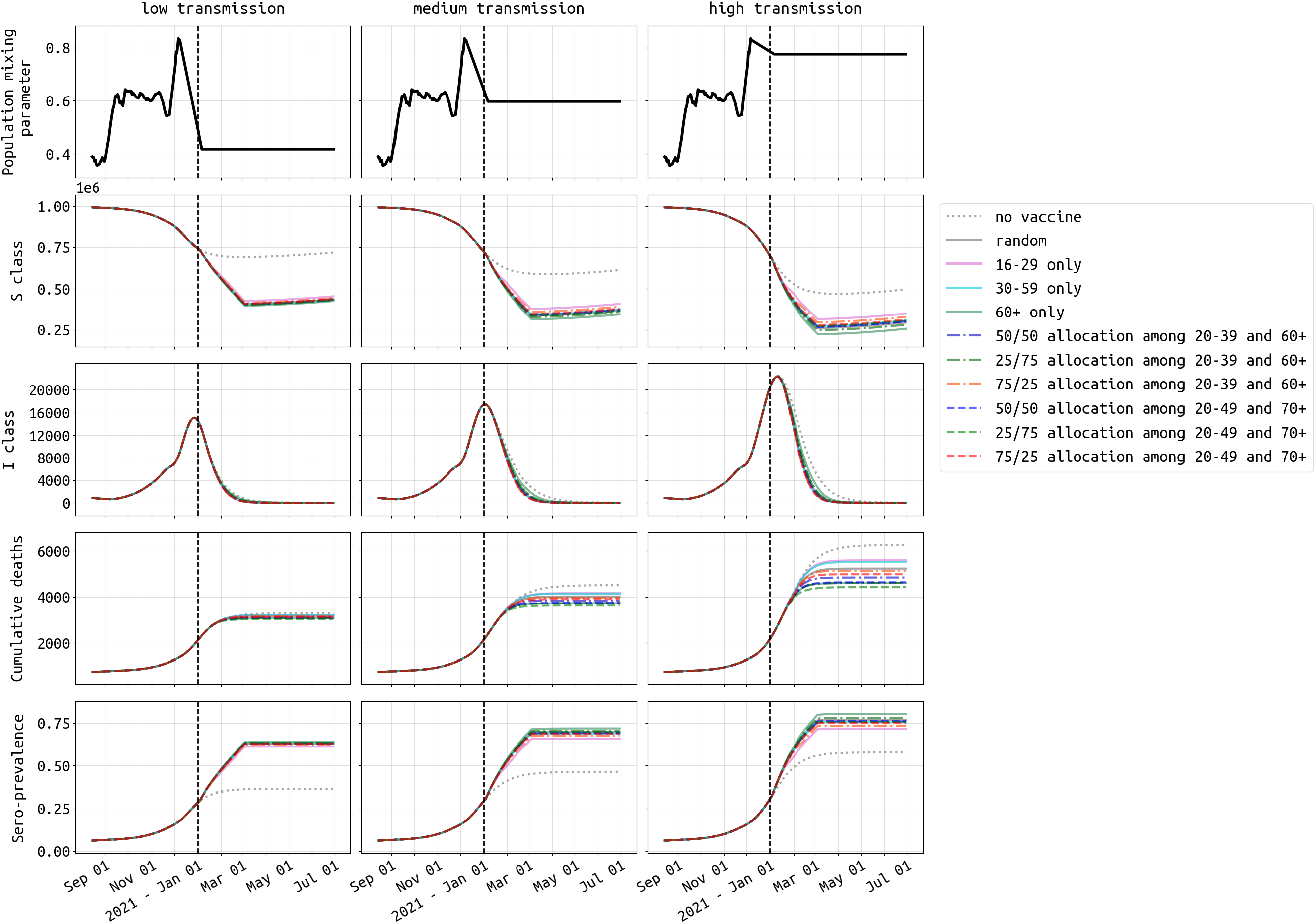
Dynamics of SARS-CoV-2 epidemic in Rhode Island under three different transmission scenarios from December 1 2020 onwards. The vaccine profile shown here has efficacy half-life of 180 days and slope of 2. The vaccination campaign covers 300, 000 people (28.3% population coverage) and ends on 04 March 2021. Under medium transmission (middle column), the population mixing parameter is assumed to remain at the mean mixing level from September through November 2020. This mixing parameter (top row) is reduced by 30% and increased by 30% to achieve low and high transmission settings, respectively. The second and third row show the size of susceptible (*S*) and symptomatic (*I*) compartment over time. The fourth row shows the cumulative deaths, both at home and in hospitals. The last row shows seroprevalence (including vaccinees) in Rhode Island from August 15, 2020 to 30 June, 2021. With no vaccination (dotted gray line), seroprevalence would reach 36.3%, 46.3%, and 57.7% by June 30, 2021 under low, medium, and high transmission settings, respectively. Scenarios and dynamics for Massachusetts shown in Figure S14.

Individuals can be vaccinated if they are in the susceptible class. Individuals who experienced asymptomatic infection and are in the recovered class can also be vaccinated, according to whether they were likely to have been asymptomatic or not [26] and confirmed PCR-positive or not (reporting parameter *ρ* in Wikle et al [17]). Individuals in the exposed class *E* or the asymptomatic class *A* can be chosen for vaccination but we assume that they progress through their normal course of disease with no effect of the vaccine, as vaccination will have occurred for these individuals at a time when their immune system has already been exposed to whole live virus SARS-CoV-2. Vaccinated individuals from *S* and *R* are moved into the vaccinated class *Z*_1_. Vaccination strategies are considered where doses are made available for 4.7% or 28.3% of the population. We call these the low and high supply scenarios: 50, 000 or 300, 000 vaccinations available in RI, and 300, 000 or 1.8 million vaccinations available in MA. Distribution lasts from 14 December 2020 through 13 January 2021 (4.7% vaccine coverage) or 14 December to 04 March 2021 (28.3% vaccine coverage). Individuals progress from *Z*_1_ through to *Z*_24_ and back to *S* over the course of 360 or 540 days.

### 2.2 Vaccination Strategies

The following vaccination strategies are considered: (1) random, where any individual *≥* 16 in the population can be chosen for vaccination on a particular day; (2) 16-29 age group only; (3) 30-59 age group only; (4) 60-and-above age group only; vaccine supply is allocated to the 20-39 and 60+ age groups in proportions of (5) 75/25, (6) 50/50, (7) 25/75; vaccine supply is allocated to the 20-49 and 70+ age groups in proportions of (8) 75/25, (9) 50/50, and (10) 25/75. If there is sufficient supply to cover an entire age group, the remaining vaccines are allocated to the second age group. If both age groups have been covered completely, the remaining vaccines are distributed at random in the population to all individuals over the age of 16.

In addition, using the nine 10-year age bands in our model, we consider all 2^9^ *−* 1 = 511 possible combinations of age bands in the vaccination strategy. We take this approach to see if including or excluding a particular age band has a large effect on the results, after marginalizing over the inclusion/exclusion of the other age groups. As evaluation criteria, we consider the total cumulative number of cases, hospitalizations, and deaths through to 30 June 2021. Our model does not evaluate the benefits of vaccinating health-care workers. We assume that health-care and front-line medical staff are vaccinated first, and that hospital capacity and staffing during the winter wave are not affected by absenteeism or a surge of COVID-19 cases.

## 3 Results

As the duration of immunity from the two current high-efficacy vaccines (Pfizer/BioNTech and Moderna) to have completed phase 3 clinical trials is not known, we build several vaccine profiles that have 95% vaccine efficacy at 60 to 90 days post-vaccination, with four different slopes of waning vaccine efficacy (Figure 2). In the most pessimistic scenario considered here, vaccine efficacy wanes to 50% after 6 months and continues to lower levels via an exponential decay thereafter (three solid lines Figure 2). Seven profiles are considered in all, and while there are quantitative differences in the population-level outcomes when considering different vaccine profiles, qualitatively the 6-month outcomes on age prioritization (see below) are not sensitive to the exact shape of the vaccine efficacy profile. Outcomes after 12 months are sensitive to the vaccine profile, with shorter half-life vaccines likely requiring booster campaigns to be planned for 2022 (results not shown).

The most straightforward approach to maximizing public health utility out of every vaccine dose is to vaccinate high-contact and high-risk individuals. It is known that contact rates vary across the age groups, and that for SARS-CoV-2 risk and clinical severity increase monotonically with increasing age. To determine the average sizes of the effects of including or excluding certain age groups for SARS-CoV-2 vaccination in Rhode Island and Massachusetts, we considered all possible strategies (511 in total) defined by inclusion/exclusion based on 10-year age band. We assumed that transmission from December 1 2020 onwards would persist at the mean level observed in Sep-Nov 2020 (the “medium transmission” scenario). The violin plots in Figure 4 show the total number of cases, hospitalizations, and deaths when a particular age group is included (blue) or excluded (orange) in a vaccination strategy. Each of the vaccination strategies considered here simply has random distribution among individuals in the included age groups. Including the middle age groups (20-39 or 20-49) results in an overall benefit in reducing case numbers, with average reductions of 1.24% (IQR: 0.94% *−* 1.64%) when including the 20-29 age group, 1.08% (IQR: 0.73% *−* 1.46%) for the 30-39 age group, and 0.34%(IQR: 0.17% *−*0.63%) for the 40-49 age group. When evaluating hospitalizations, including the 70-79 age group results in 0.55% (IQR: 0.43% *−* 0.72%) fewer hospitalizations, and including the 80+ age group results in 1.06% (IQR: 0.84% *−* 1.35%) fewer hospitalizations. When evaluating deaths as the relevant outcome measure, including the 70-79 age group results in 0.67% (IQR: 0.48% *−* 0.95%) fewer deaths, and including the 80+ age group results in 3.95% (IQR: 3.28% *−* 5.08%) fewer deaths. As expected, the high-risk (70+) and high-contact (20-49) age groups should be priority targets for vaccination campaigns, with an elderly focused campaign being the simplest approach to minimizing fatalities in the short-term.

**Figure 4:**
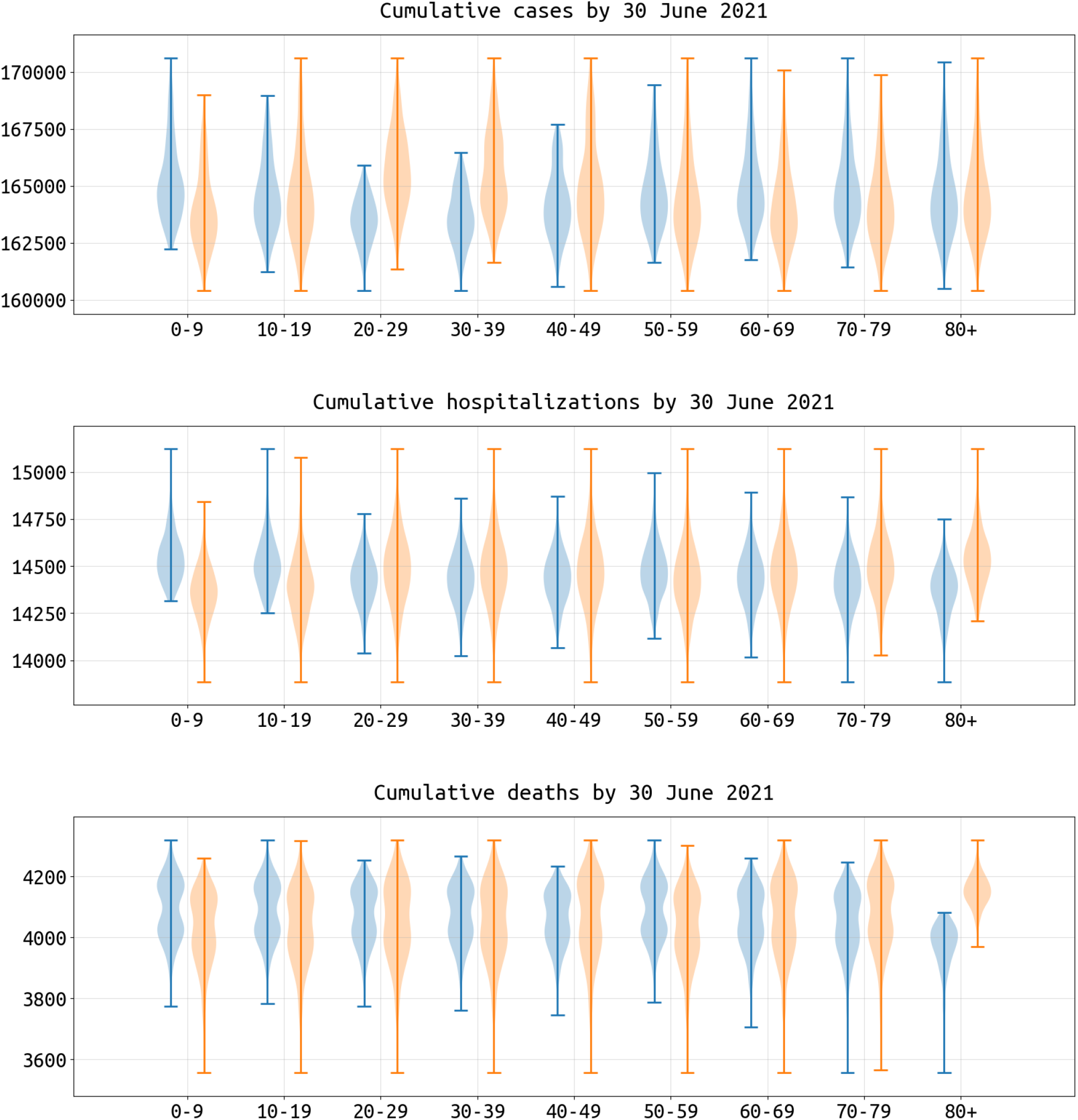
Impact of including (blue) or excluding (orange) each age group in a vaccination policy, measured as reductions in cumulative cases, hospitalizations, and deaths in Rhode Island by 30 June 2021. With nine age classes in the model, there are 2^9^ *−* 1 = 511 possible age-based vaccination strategies where the vaccine supply is equally distributed among the participating age groups. The vaccination campaign in this figure ends on 4 March 2021 and the total vaccine supply is enough to vaccinate 300, 000 people (28.3% coverage). Vaccine efficacy half-life here is 360 with slope = 2. Each violin plot shows the distribution across 256 (blue) or 255 (orange) strategies which include or exclude the corresponding age group. Campaigns which cover the 20-29 age group would reduce the median of cumulative cases by 1.24% and cumulative hospitalizations by 0.38% compared to those not covering this age group. The median of cumulative deaths drops by 3.95% when targeting the 80+ age group when compared to strategies that do not include the 80+ age group. Results for Massachusetts shown in Figure S13.

In a low-supply scenario with 50, 000 vaccinations available in Rhode Island (4.7% coverage) during the initial rollout, allocating 25% of vaccines to the 20-49 age group and the remaining 75% to the 70+ age group is optimal (among the strategies evaluated) in terms of minimizing deaths and hospitalizations; see left three columns, Figure 5. While the two different 25/75 (younger/older) allocations we evaluated are optimal for death and hospitalization outcomes, they are associated with the highest final case counts meaning that they have the smallest effect on overall transmission reduction. The 75/25 allocations are best at reducing case counts and near-optimal at reducing hospitalizations (red lines, Figure 5). But, as the 75/25 allocations are majority focused on the younger age classes, they are associated with a substantially higher final death count by mid-2021 as they fail to provide enough vaccination for the age groups with the highest risk of dying if infected. Figure 6 (top row) shows that prioritization of vaccine allocation to the 70+ age group has a modest effect on reducing hospitalizations and a substantial effect on reducing deaths, as much as a 7% difference in cumulative deaths through June 30 2021 when viewing the two extreme strategies (10/90 versus 90/10 allocations). This corresponds to more than a hundred deaths in RI and hundreds of deaths in Massachusetts. Because the vaccine supply is low absolute benefits are also low, and most age-allocation strategies are associated with case/hospitalization/death outcomes that are within 5% of a simple random allocation strategy.

**Figure 5:**
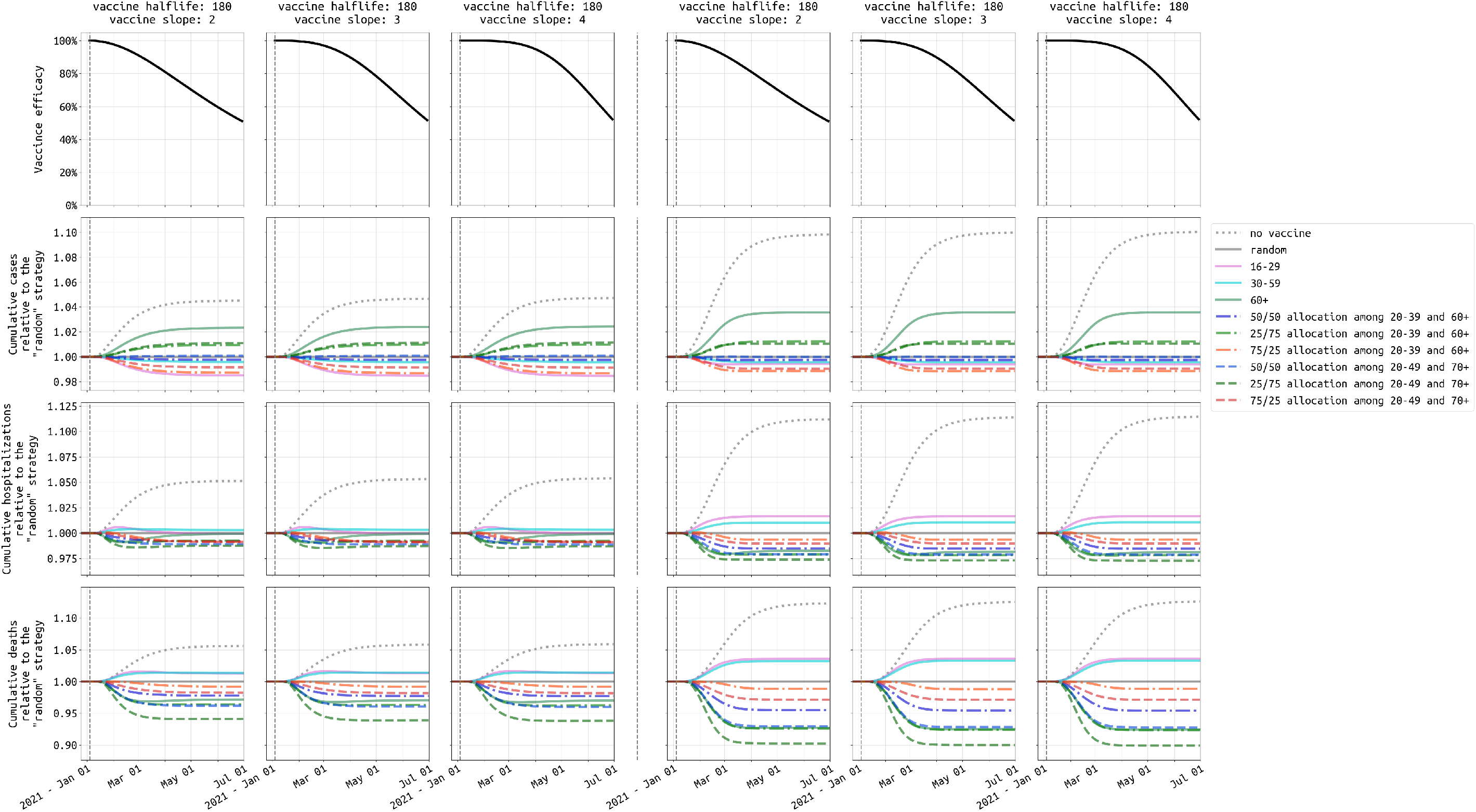
Comparison of ten vaccination strategies in Rhode Island based on cumulative cases (second row), hospitalizations (third row), and deaths (fourth row), given two levels of vaccine supply and under the three most pessimistic assumptions for vaccine profile (top row). The left three columns show outcomes when the vaccine supply is low (50,000 vaccines), and the right three columns show outcomes for high vaccine supply (300,000 vaccines). Our reference strategy is “random” (solid gray line) where everybody has an equal chance of getting vaccinated. The solid pink, cyan, and green lines are strategies which cover the 16-29, 30-59, and 60+ age groups, respectively. The dashed-dotted lines are strategies targeting 20-39 and 60+ age groups with different distribution of vaccine supply to the two age groups (e.g. blue is 25% of the allocation for 20-39 and 75% of the allocation for 60+). The dashed lines are strategies which include 20-49 and 70+ age groups, which are the key age groups identified in Figure 4. Results for medium transmission scenario in Massachusetts shown in Figure S20.

**Figure 6:**
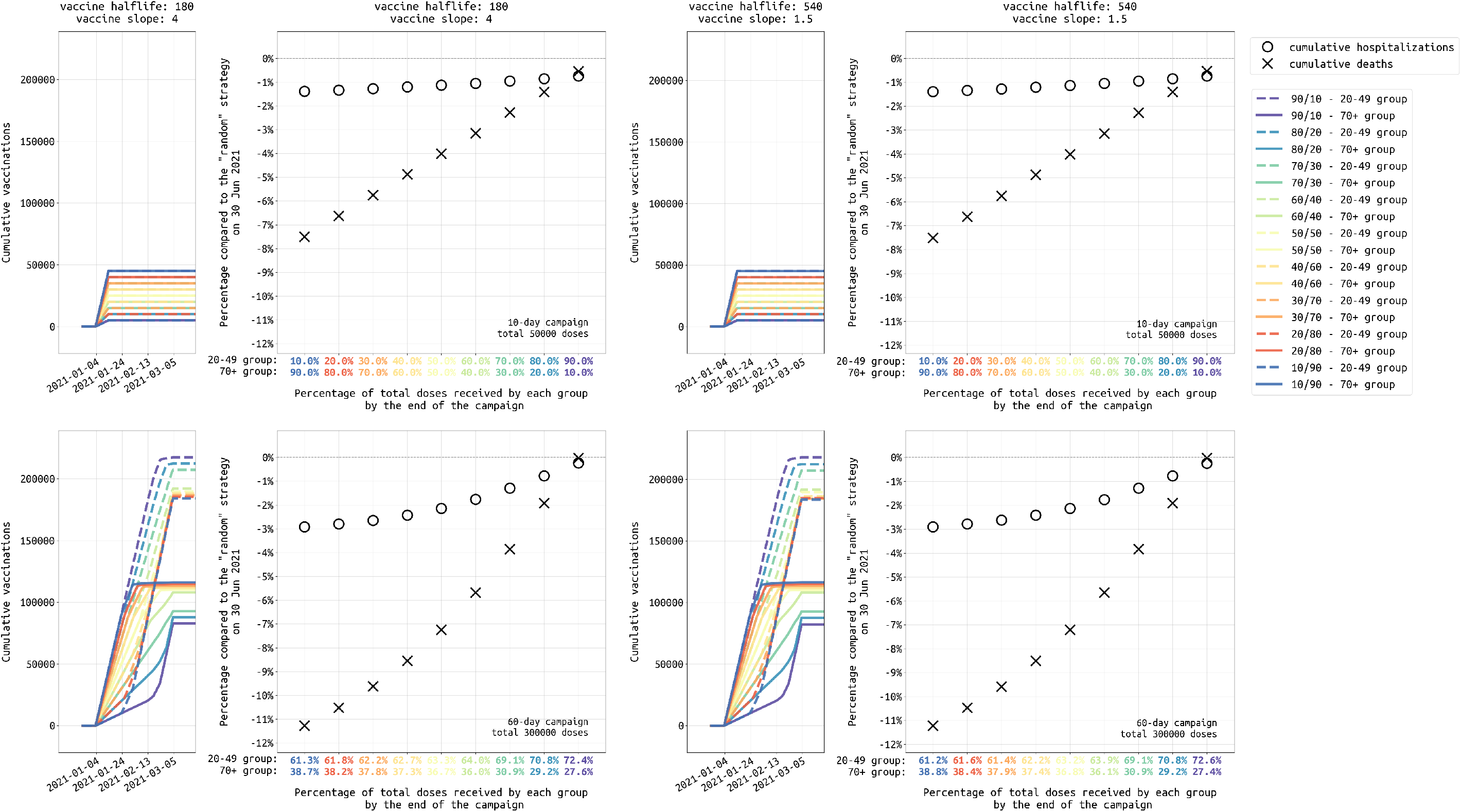
Analysis of different age allocations for strategies focused on the 20-49 and 70+ age groups, ranging from a 10/90 allocation (90% of vaccines initially given to 70+ age group) to a 90/10 allocation (90% of vaccines initially given to 20-49 age group). Top row shows the low supply scenario for Rhode Island (50,000 vaccines) and bottom row shows the high supply scenario (300,000 vaccines available). Left and right columns show scenarios for different vaccine profiles, with a 180-day half-life vaccine shown on the left and a 540-day half-life vaccine on the right. Narrow plots show the cumulative vaccinations by age class, for the 20-49 (dashed lines) and 70+ (solid lines) groups under different allocation strategies. Circles and crosses show the percentage reduction in hospitalizations (circles) and deaths (crosses) when compared to a random distribution strategy. The *x*-axis shows the final vaccine allocation to the 20-49 and 70+ age groups. In the top row (low supply), these allocations are 10/90, 20/80 etc., as planned. But, in the bottom row, there is nearly enough vaccine for all individuals in these two age classes, and when one age group is fully vaccinated the remainder of the supply is used for the second age group. Thus, while the final age distribution does not vary much (61/39 to 73/27) under amply vaccine supply, it does affect which groups get vaccinated earlier. Under ample vaccine supply, a 10/90 allocation means that older individuals get vaccinated earlier and more deaths and hospitalizations are averted. Results for medium transmission scenario in Massachusetts shown in Figure S23.

In a more optimistic scenario — 300, 000 vaccines procured in Rhode Island (28.3% population coverage) — strategies focused on the elderly can reduce cumulative death numbers by as much as 10% when compared to a random distribution strategy (three right columns, Figure 5). The optimal strategy among those evaluated is a 25/75 distribution to the 20-49 and 70+ age groups, outperforming the 60+ strategy and the 20-39 and 60+ strategy with 25/75 allocation, both of which vaccinate more elderly individuals but have sub-optimal outcomes because they have too small of an effect on transmission reduction. With ample vaccine supply, most individuals in the 20-49 and 70+ age groups will be vaccinated. However, the allocation is still important in deciding which groups are vaccinated earlier than others. Figure 6 (bottom row) shows that the percentage of the entire allocation going to the 20-49 and 70+ age groups does not differ greatly between a starting allocation of 90/10 and 10/90 (because everyone is eventually vaccinated anyway), but in a 10/90 allocation (i.e. 90% of vaccines reserved for 70+) the older age groups receive the vaccine earlier. Under a 10/90 allocation, the mortality benefits are substantial, with 11% fewer deaths when compared to a 90/10 allocation in which the 20-49 age group is vaccinated earlier. For both low and high supply scenarios, strategies focusing predominantly on higher-contact age groups – e.g. strategies where 50% or more of the vaccine supply is allocated to younger age groups – are always sub-optimal at minimizing deaths.

The high and low vaccine supply scenarios have identical implications for the current epidemiological situation in Massachusetts. Figure S20 shows that the 25/75 vaccine allocation to the 20-49 and 70+ age groups is also the optimal allocation (when considering deaths as the primary outcome) among those examined for Massachusetts. Under a scenario of low vaccine supply, deaths are reduced by approximately 8% when comparing to a strategy of random vaccine allocation; under high vaccine supply deaths are reduced by approximately 12% when comparing to a random strategy. As in Rhode Island, prioritization of the 70+ group (or any group with a similar mortality risk) leads to optimal outcomes (see Figure S23).

For Rhode Island, assuming a total of 300, 000 vaccinations (28.3% coverage), a strategy that is targeted 25*/*75 at the 20-49 and 70+ age groups will results in 375 fewer hospitalizations and 393 fewer deaths, by June 30 2021, than a random allocation strategy. Compared to the unmitigated scenario (no vaccine), this vaccination strategy will save 887 more lives and reduce hospitalization numbers by 1978 by the end of June 2021. In Massachusetts, with an ample supply of vaccines (1.8M vaccines distributed by March), this same approach translated to 1222 fewer hospitalizations and 3007 fewer deaths compared to a random strategy, and a total of 15, 578 fewer hospitalizations and 7502 fewer deaths compared to a scenario with no vaccine. Our current seroprevalence estimates indicate that when the distribution campaign is finished on 4 March 2021, the population-level immunity (combined natural and vaccine-induced) in Rhode Island will be 71.3% and in Massachusetts will be 61.1%.

Vaccination of seropositive individuals can lead to wastage of vaccines, especially if the seropositive individuals were infected recently. Figure 7 shows that a policy of vaccinating only seronegative individuals would result in 1% to 2% fewer hospitalizations and deaths through mid-2021 than policy where serostatus is not checked prior to vaccination.

**Figure 7:**
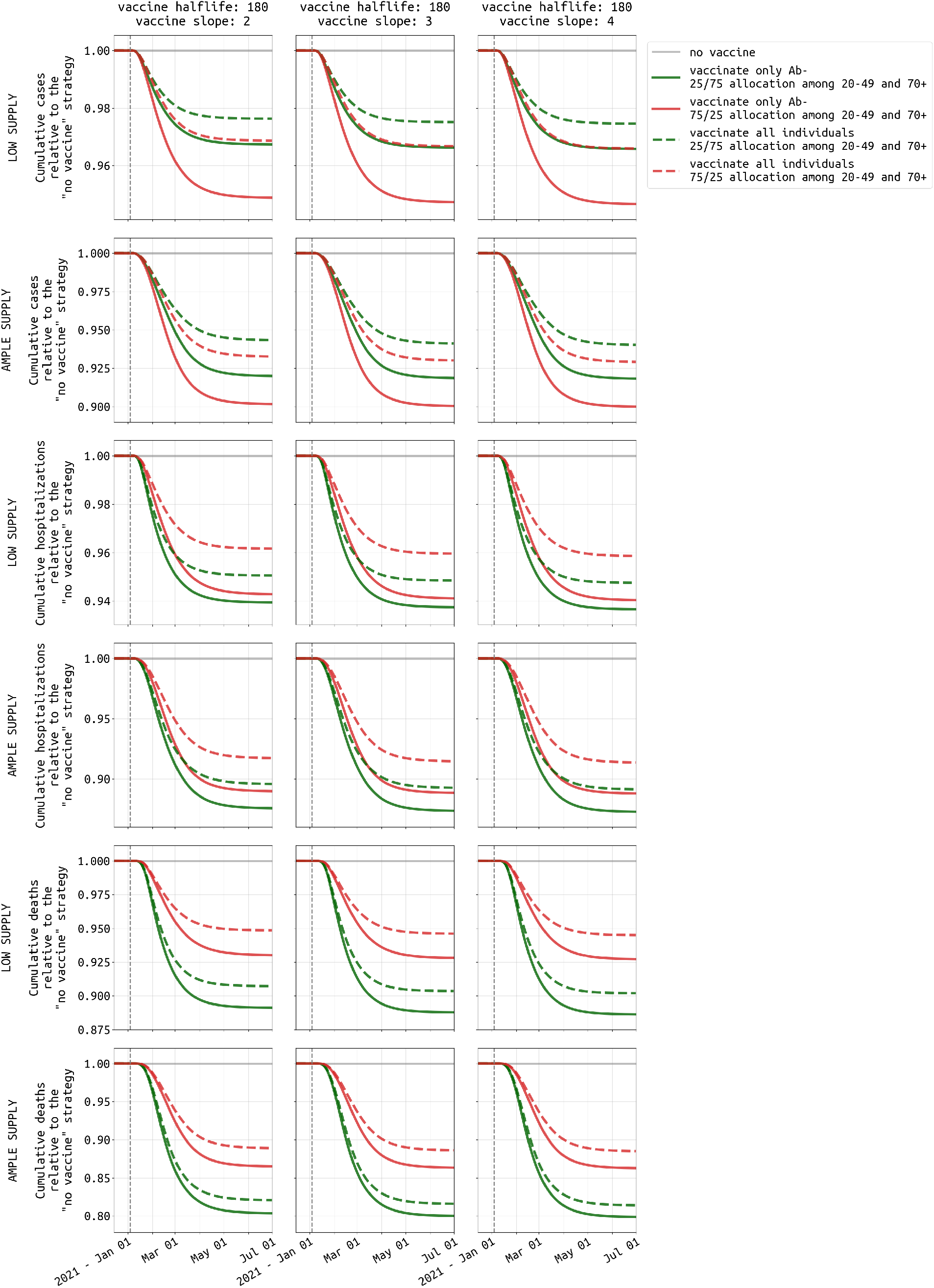
Difference between vaccinating all individuals (dashed lines) and vaccinating only antibody-negative individuals (solid lines). Three different vaccine profiles are considered (three columns). Plots show cumulatve cases (rows 1 and 2), hospitalizations (rows 3 and 4), and deaths (rows 5 and 6) under both low supply (50,000 vaccines available) and ample supply (300,000 vaccines available). Green lines show the preferred 25/75 allocation in the 20-49 and 70+ age groups, and red lines show a 75/25 vaccine allocation. In Rhode Island, a vaccination campaign that is able to vaccinate only antibody-negative individuals will result in 1% to 2% fewer hospitalizations and deaths. Results for Massachusetts shown in Figure S24.

## 4 Discussion

Two key characteristics of epidemic management that we account for when devising optimal vaccine distributions are (1) current seroprevalence levels in Rhode Island and Massachusetts and (2) the waning effect of vaccine efficacy. End-of-2020 attack rate is expected to be between 26% and 32% in MA and RI, approximately, assuming that November trends held through December. These levels are too low to hope that vaccination campaigns will push populations past the point of herd immunity in the first two months of 2021. Nevertheless, assuming vaccine supplies can reach 20% to 30% coverage statewide by spring, both Rhode Island and Massachusetts may reach approximately 60% to 70% population immunity by spring 2021, substantially slowing the spread of the virus by late spring and summer. These approximations assume our medium transmission scenario, and do not yet account for the arrival and spread of the higher-transmission B.1.1.7 lineage identified in the UK in late December [27, 28]. Vaccine efficacy is unlikely to wane so quickly that large groups of vaccinated individuals will be at risk of reinfection in mid-2021, however a small proportion of individuals infected in March-August 2020 may be at risk of reinfection if their antibody levels were to wane to sub-protective levels one year post-infection. The key variable to keep track of in summer/fall 2021 will be duration of immunity — both natural and vaccine-induced — to understand if the population-level risk of renewed outbreaks is likely to return in fall 2021. If duration of immunity is short, especially in the older age groups, booster vaccinations may be needed in late 2021 or in 2022.

The basic outcome observed in our evaluations is that high-mortality groups need to be vaccinated first in order to minimize death counts. In our model, groups that are at high risk of death if infected are the older age groups, but our analysis implies that any high-risk group — whether the risk factor is age, obesity, diabetes, past lung disease, lack of health care access, or anything else — should have equally high priority to vaccination. The obvious second-order implication is that individuals who have high contact rates with risk groups or cohabit with someone at elevated risk of death should similarly be prioritized for vaccination. Although the highly specific context in which we have been managing COVID-19 risk over the past year will allow us to identify such individuals on a case by case basis, there does not seem to be a systematic way to define this second-order group who have either frequent contacts or cohabitation with high-risk individuals; the clear exceptions are health-care workers and employees of long-term care facilities.

Vaccination of high-contact groups alone is not an optimal approach, primarily due to the mortality differences between the middle and the older age groups. This conclusion is identical to the one reached in Bubar et al [14], but it differs from many common optimal vaccine allocations for influenza virus where distribution to high-contact groups can be optimal under a wide range of conditions [11–13].

One efficiency to capture in the early part of the vaccination campaign — especially when supply is moderate — is the deprioritization of vaccination for individuals with past confirmed COVID-19 infections or recent confirmed COVID-19 infections. Although this is currently voluntary, public health communication around this topic could ask individuals with known past infection to offer up their place in the vaccination queue to those that are still fully susceptible. In Rhode Island and Massachusetts, more than 75% of symptomatic COVID-19 cases were likely identified through testing during 2020, meaning that a large majority of individuals would be aware that they had a past infection. Asymptomatically infected individuals will not know that they were COVID-positive at some point in 2020, unless they were included in a random screening campaign focused on nursing home staff/residents or essential workers. Most asymptomatic infections during 2020 would have occurred in the *<* 20 or *<* 30 age groups, individuals who would not be prioritized for vaccination, thus reducing the effect that serostatus would have on vaccine wastage in the early part of the campaign. Given the pace and potential for delays during the initial roll-out and delivery of vaccines, it is unlikely that serological testing (prior to vaccination) would be of any benefit to the campaign overall, and it would likely add to delays unless rapid high-specificity point-of-care IgG tests were used. Our model currently assumes that individuals with past known infection will not be part of the winter vaccination campaign; if these individuals are included, the hospitalization and mortality benefits of the vaccination campaign will be lower by about 1% or 2%.

The major benefit in focusing on state-level analyses of vaccine roll-out is that cumulative seroprevalence and vaccination numbers can be tracked on a month by month basis. This will be critical for public communication in the coming 4-6 months as the public will be eager to know whether total infection and vaccination counts are in the range of “30+10” or “40+20”, allowing the public health system to describe risk in terms of the fraction of individuals that are still not immune. Assuming current infection trends hold and vaccination trends improve to approximately 0.5% of the population per day, both Massachusetts and Rhode Island should cross the 50% immune mark in February or March 2021. Two possible paths are possible after this point. If the more transmissible B.1.1.7 lineage has spread to New England and begun to dominate infections in RI and MA, we will need to wait as vaccination/immunity numbers increase since the new variant will almost certainly be associated with a higher herd-immunity threshold. If the epidemics in RI/MA wind down without the introduction of B.1.1.7, restrictions on gathering sizes, school cohorting, masking, travel, and business openings will be able to be gradually lifted as the risk of infection drops while the population progress from 50% to 70% immunity.

### Limitations

Our modeling approach has several limitations. First, although age stratification allows for a straight-forward strategy design focused on protecting the elderly, our model does not include any variables on race, comorbidities, health care workers, or other essential workers. This means that some key high-contact and high-risk groups are omitted from the modeling, and it is unlikely that an age-group proxy would be a suitable substitute for any of them. For example, health care workers would likely fall into the high-contact category, but they may preferentially be in contact with non-susceptibles (i.e. SARS-CoV-2 positives) making them more a high-exposure group than a high-contact group. In addition, the benefits of vaccinating HCWs and essential workers is that certain essential services (hospitals, schools, grocery stores) can continue functioning, a benefit not captured in traditional epidemiological models.

Second, Phase 3 efficacy trials of the Pfizer/BioNTech and Moderna vaccines were not designed to evaluate reductions in transmission. Thus it is not possible to state whether the vaccines’ high efficacy could be compromised by asymptomatic or sub-clinical infections occurring in vaccinated individuals and allowing for the continuation of transmission. Asymptomatic individuals do have lower viral loads and fewer opportunities to transmit via large droplets projected out through coughs, sneezes, speech, or breathing. Therefore, inadequate vaccine prevention of potentially-transmissible asymptomatic infections may reduce the indirect benefits of vaccination, but this effect size is currently unknown (and possibly small). Asymptomatic and presymptomatic infection does occur for SARS-CoV-2, but the naturally observed infectivities in household and contact tracing contexts are likely to be different than the infectivity of a vaccinated infected asymptomatic individual. Preliminary reports from a Moderna FDA filing [29] have begun to be interpreted as early evidence that the Moderna vaccine may offer some degree of protection against asymptomatic infection. Understandably, this is a difficult outcome to measure in a standard vaccine trial as the frequency of planned molecular-diagnostic follow-ups may not be powered to catch a large number asymptomatic infections. In our model analysis, if vaccinated individuals were added to a ‘partially susceptible pool’ and allowed to be infected with mild or no clinical symptoms, case numbers would increase but the hospitalization and death outcomes in our analysis are unlikely to be affected.

Third, results on long-term efficacy and efficacy by age are currently unknown. The combination of these two is critical as older individuals may be protected for a shorter time than younger healthier individuals. If vaccine-induced immunity were to wane quickly in older individuals, the vaccine would be least effective in the group that needs it the most. All vaccination campaigns would then need to be restructured as routine/repeat campaigns that focus on the most vulnerable individuals and account for the fact that revaccination may need to occur often to realize the vaccination campaign’s intended mortality benefits.

### Conclusion

Real-time knowledge of seroprevalence can guide decisions on vaccine allocation. Knowing which population groups have experienced the most infection and how far seroprevalence has advanced population-wide can help determine the total vaccine supply needed as well as its distribution priorities. This highlights the value of (1) real-time attack-rate estimation, (2) completeness of population surveillance, and (3) widespread testing. Comprehensive surveillance and widespread testing ensure that individuals are as informed as possible about their sero-status to SARS-CoV-2 and allows for the health system to communicate to seropositive individuals that they should forego vaccination in the first few rounds of allocation while immunologically naive individuals are vaccinated first. Given current seroprevalence levels in Rhode Island and Massachusetts, older age groups and vulnerable populations with substantial risk of hospitalization or death resulting from SARS-CoV-2 infection should be prioritized for vaccination, after the completion of vaccine rollout for frontline health and medical workers.

## Research Support

MFB, TNAT are funded by a grant from the Bill and Melinda Gates Foundation (INV-005517). FY is supported by the NIH/NIAID Center of Excellence in Influenza Research and Surveillance contract HHS N272201400007C. KB was partially supported by the National Institute of General Medical Sciences of the National Institutes of Health under award number R35GM133700. WPH is funded by an award from the NIGMS (U54 GM088558). JA is funded by the Penn State MRSEC, Center for Nanoscale Science, NSF DMR-1420620. EH was partially supported by NSF DMS-2015273.

## Data Availability

All data and code are available at https://github.com/bonilab/covid19-vaccine-allocation-RI-MA

## Data and Code Availability

All data and code are available at https://github.com/bonilab/covid19-vaccine-allocation-RI-MA.

