## Supplementary Materials for "Optimal SARS-CoV-2 vaccine allocation using real-time seroprevalence estimates in Rhode Island and Massachusetts"

**Supplementary Material for** "Optimal SARS-CoV-2 vaccine distribution using real-time sereprevalence estimates in Rhode Island and Massachusetts"

Tran, Wikle, Albert, et al.

Date of this draft: January 10, 2021.

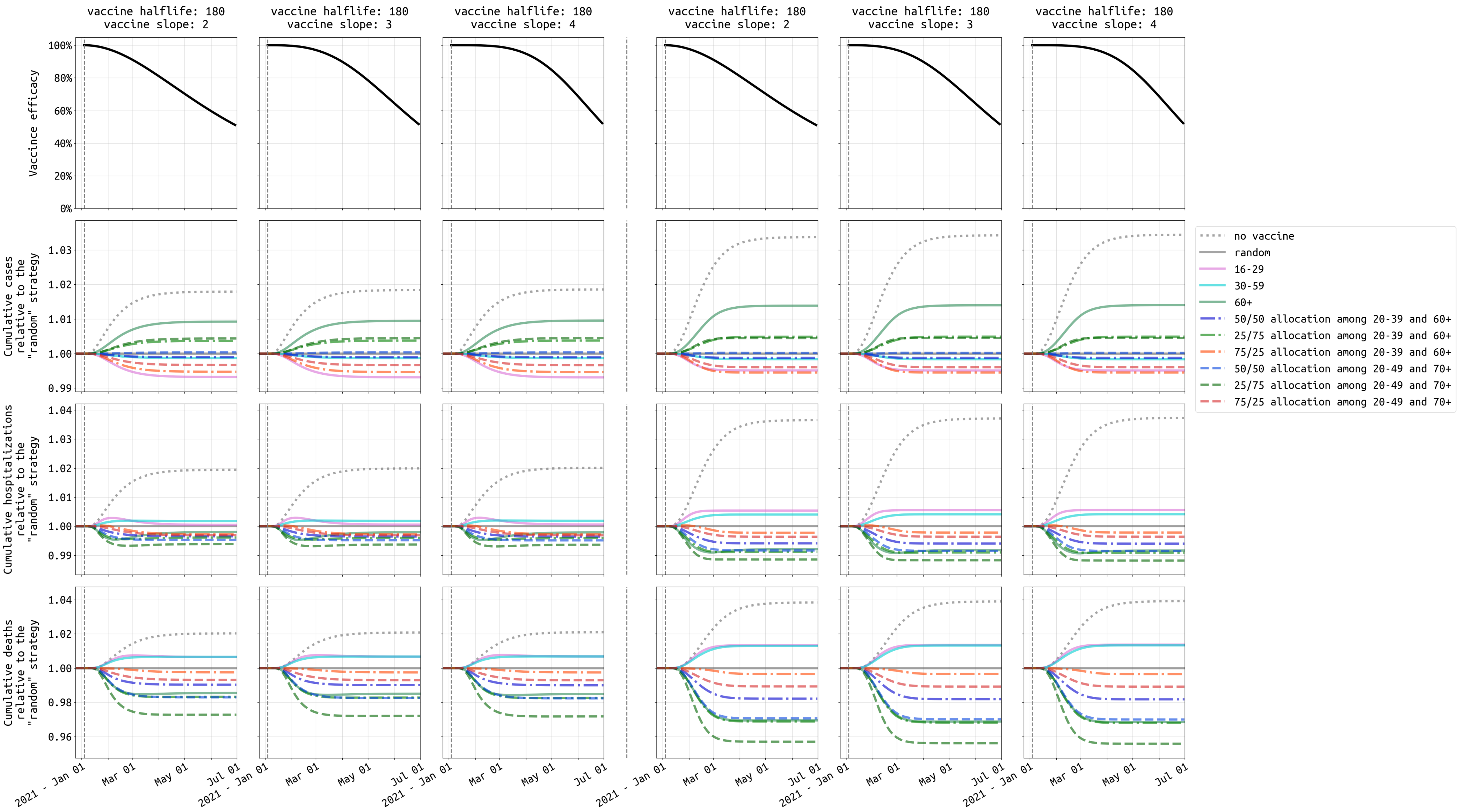


Figure S1: Similar to Figure 5, comparison of ten vaccination strategies with vaccine efficacy half-life of **180** days under **low** transmission setting in **Rhode Island**.

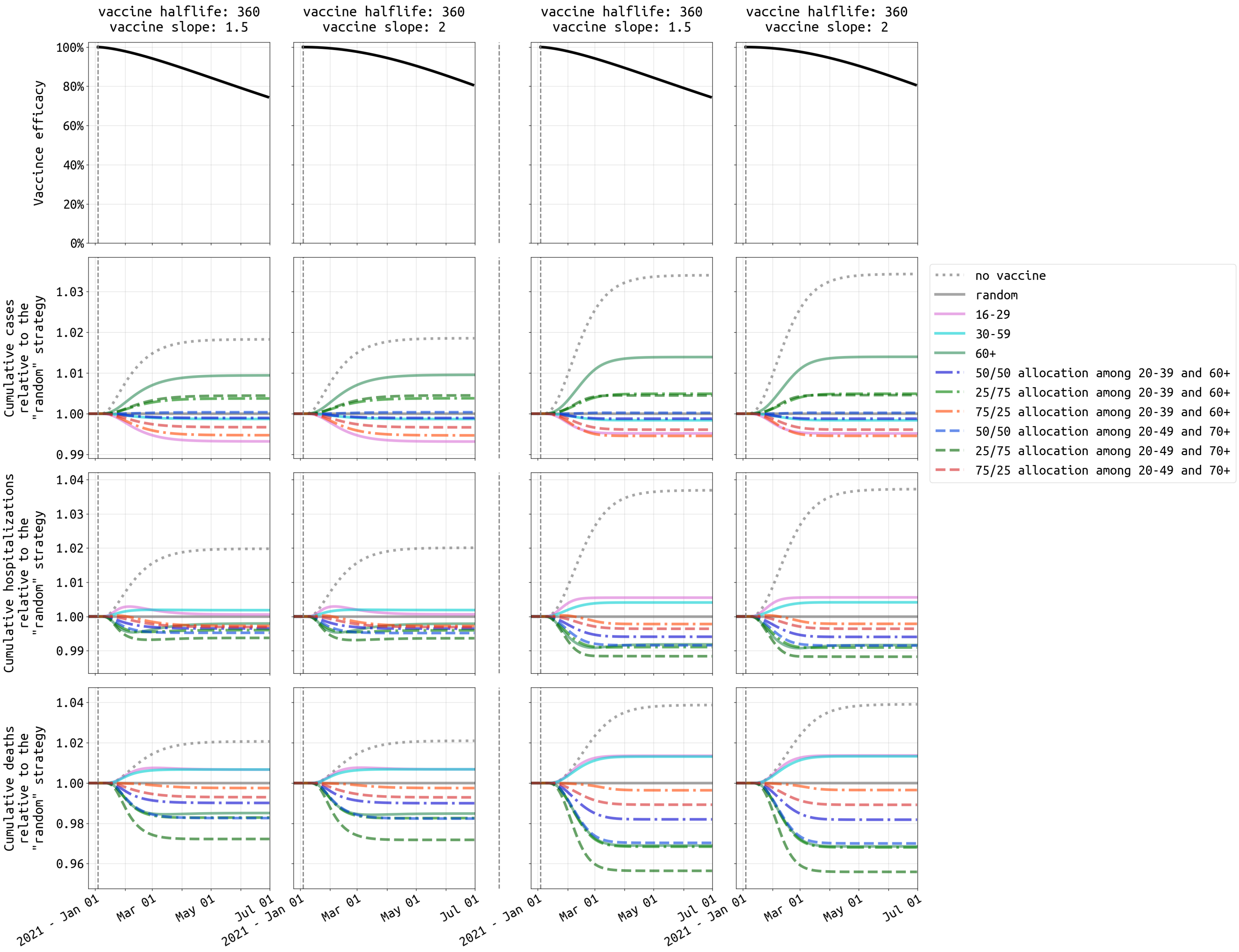


Figure S2: Similar to Figure 5, comparison of ten vaccination strategies with vaccine efficacy half-life of **360** days under **low** transmission setting in **Rhode Island**.

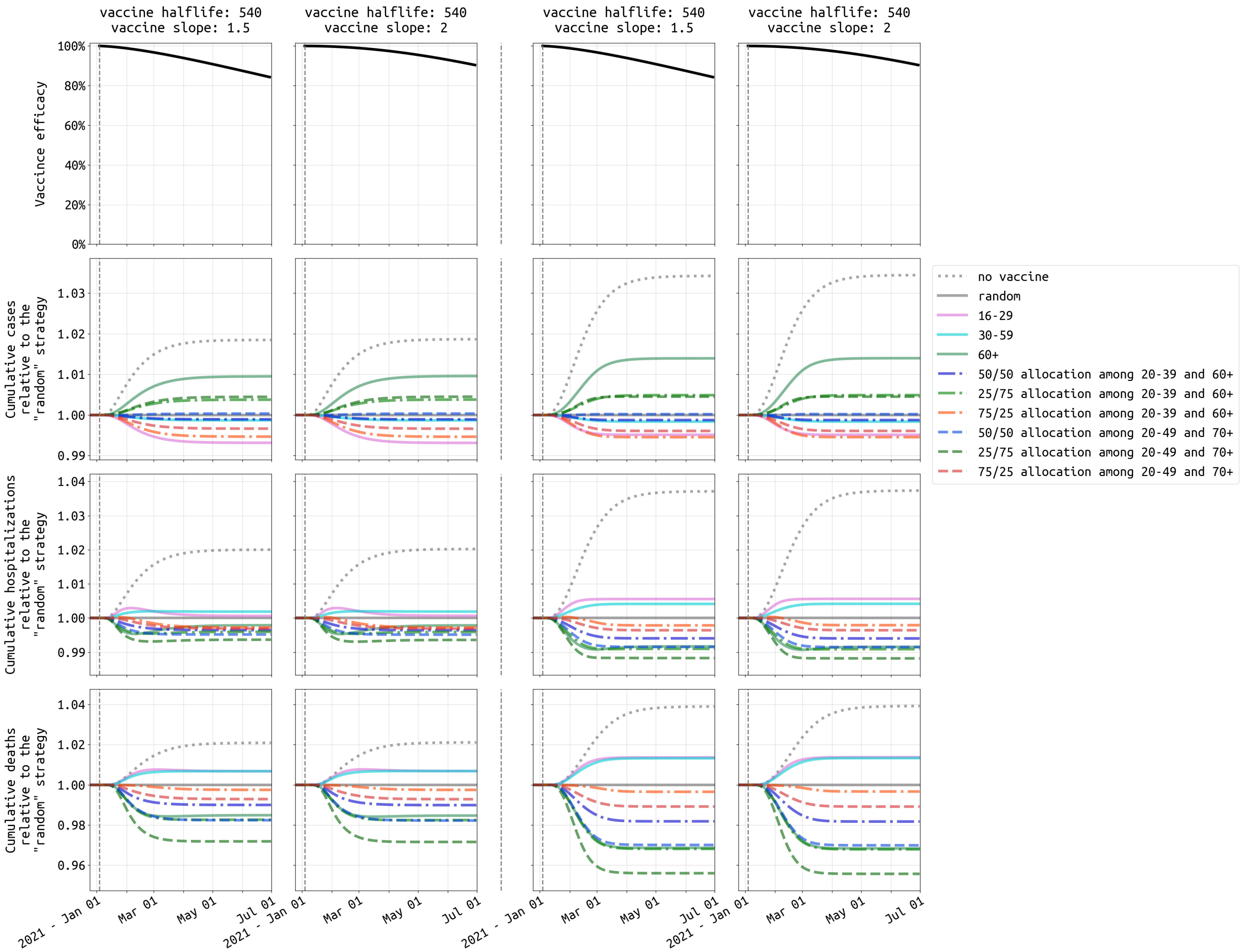


Figure S3: Similar to Figure 5, comparison of ten vaccination strategies with vaccine efficacy half-life of **540** days under **low** transmission setting in **Rhode Island**.

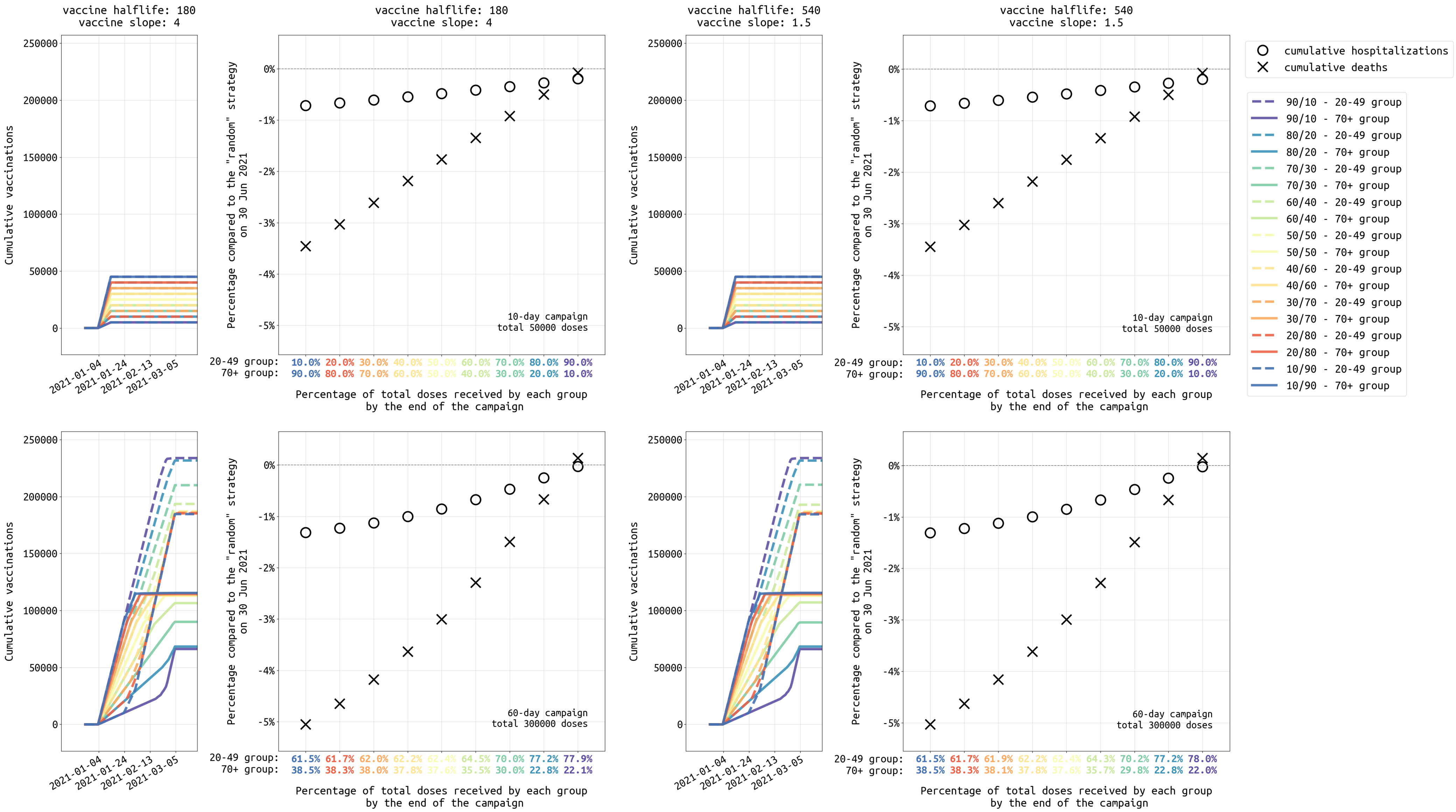


Figure S4: Similar to Figure 6, analysis of different dose allocations for strategies focused on the 20-49 and 70+ age groups, ranging from a 10/90 allocation (90% of vaccines initially given to 70+ age group) to a 90/10 allocation (90% of vaccines initially given to 20-49 age group) under **low** transmission setting in **Rhode Island**.

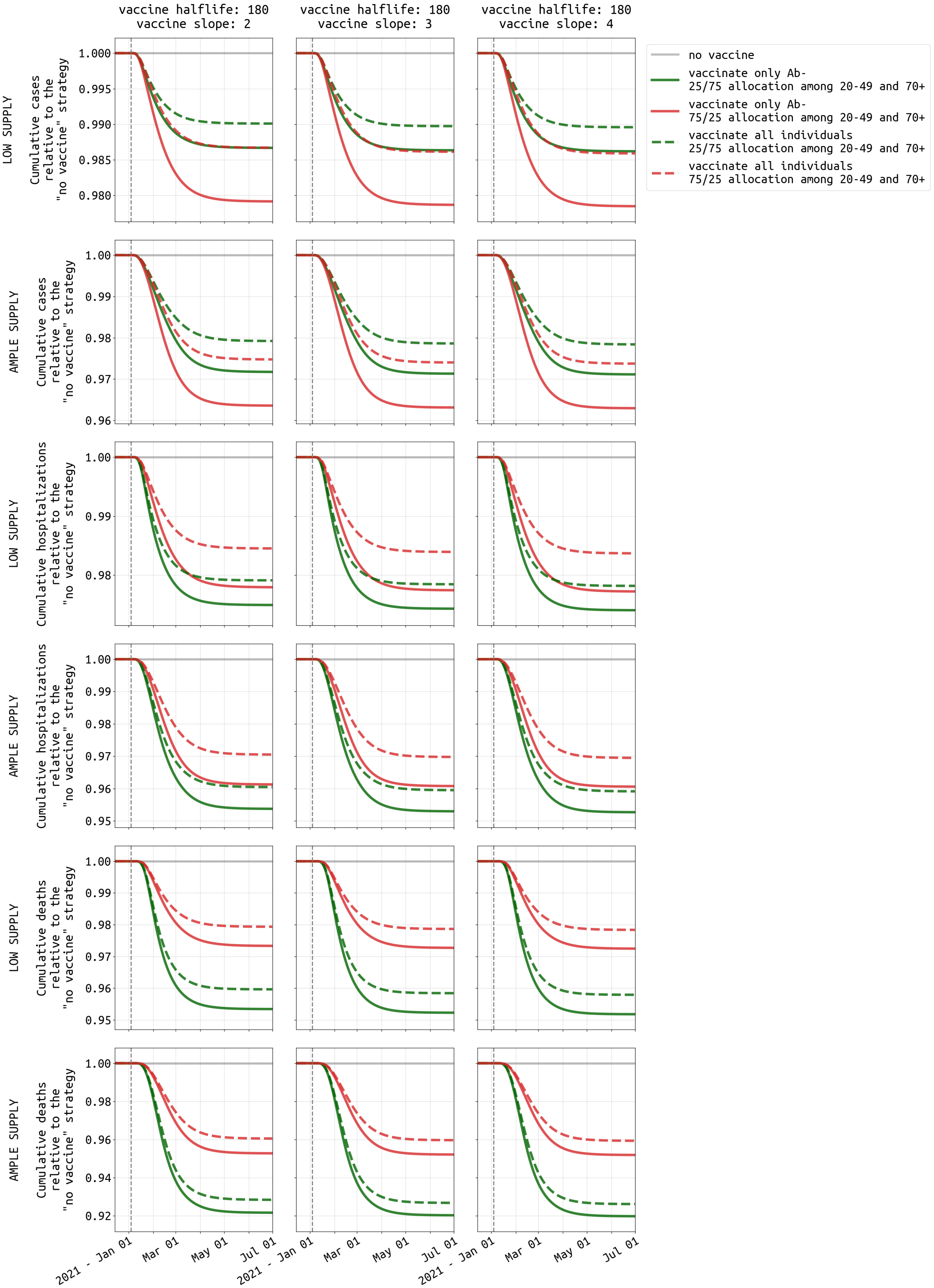


Figure S5: Similar to Figure 7, comparison of vaccinating all individuals and vaccinating only antibody-negative individuals under **low** transmission setting in **Rhode Island**.

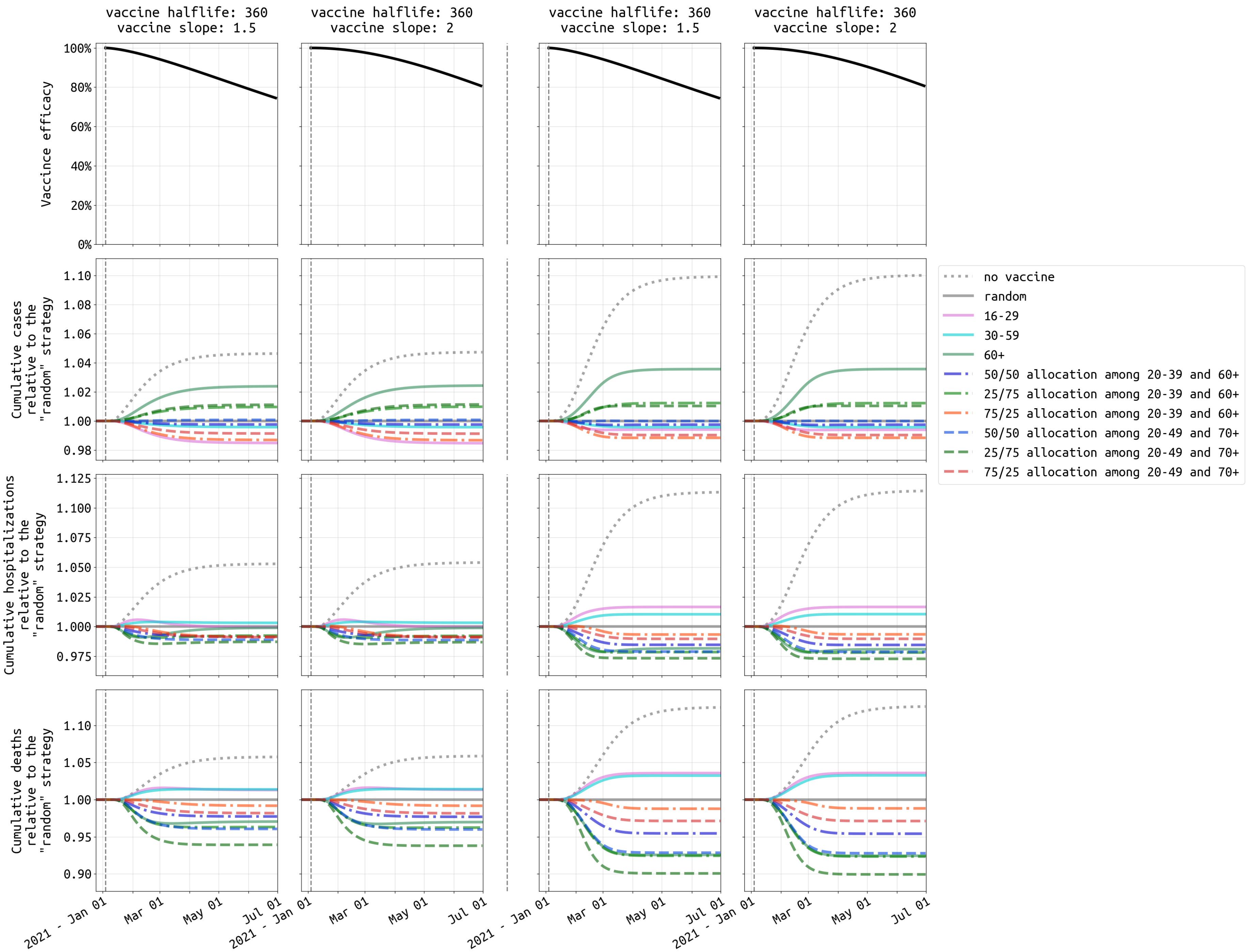


Figure S6: Similar to Figure 5, comparison of ten vaccination strategies with vaccine efficacy half-life of **360** days under **medium** transmission setting in **Rhode Island**.

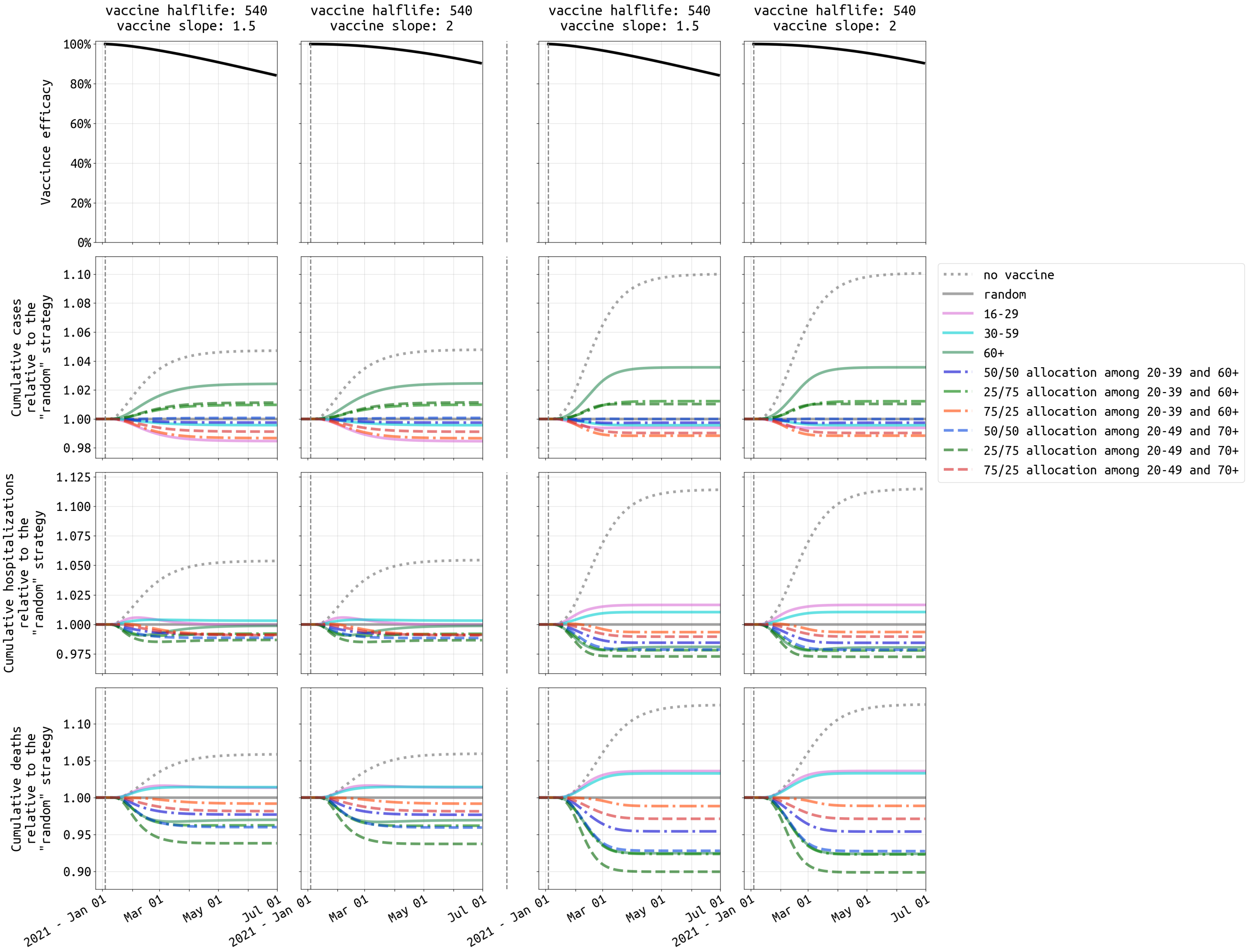


Figure S7: Similar to Figure 5, comparison of ten vaccination strategies with vaccine efficacy half-life of **540** days under **medium** transmission setting in **Rhode Island**.

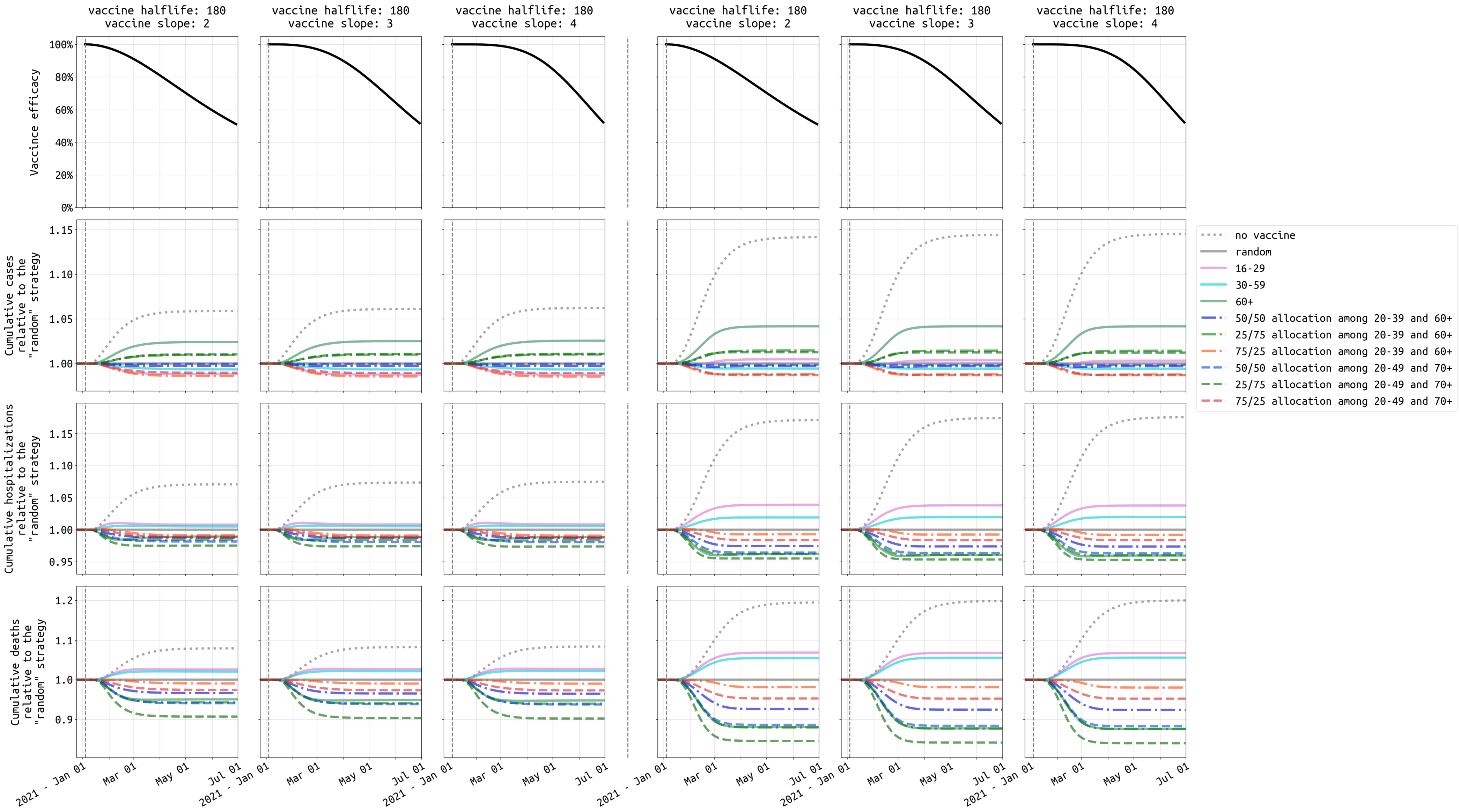


Figure S8: Similar to Figure 5, comparison of ten vaccination strategies with vaccine efficacy half-life of **180** days under **high** transmission setting in **Rhode Island**.

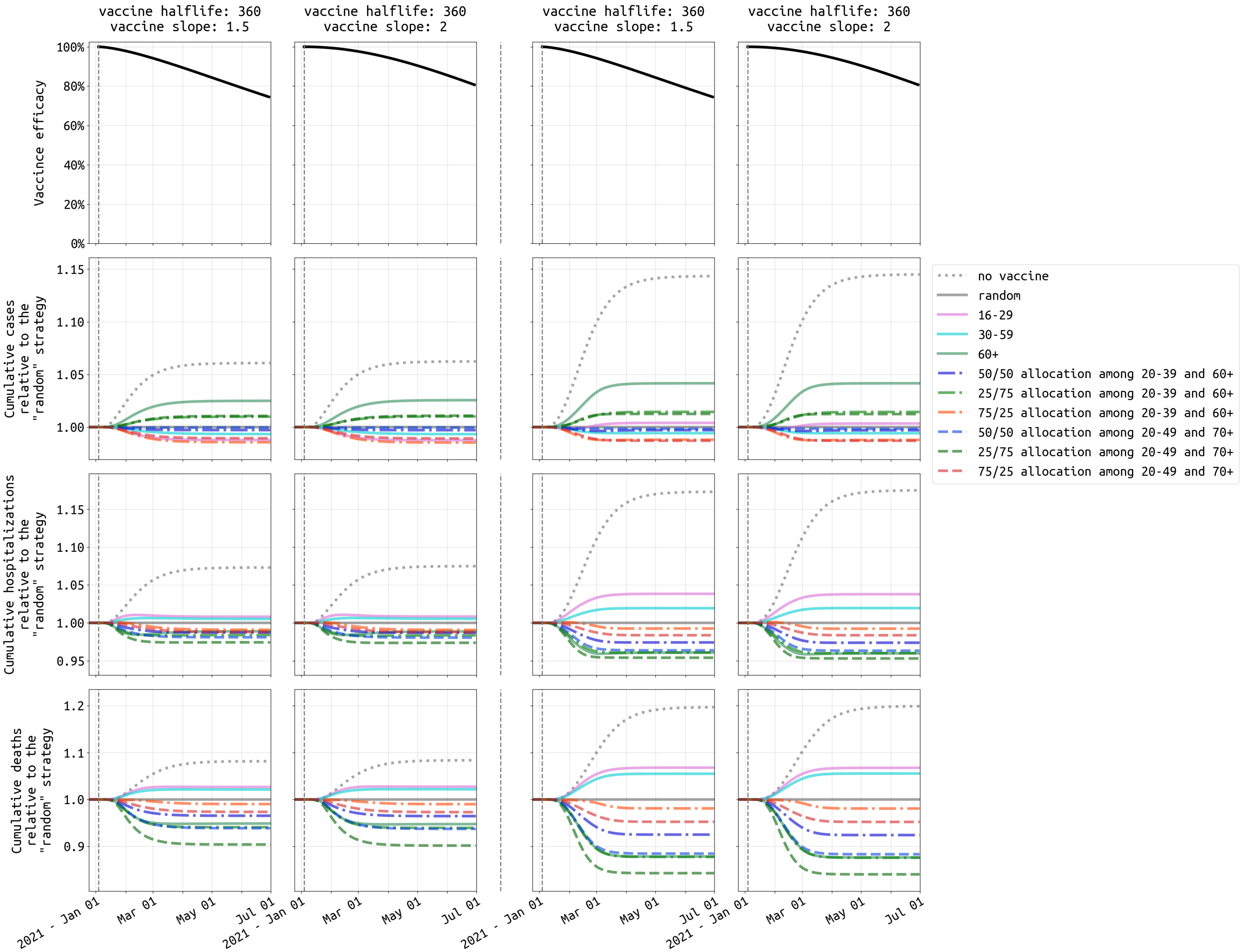


Figure S9: Similar to Figure 5, comparison of ten vaccination strategies with vaccine efficacy half-life of **360** days under **high** transmission setting in **Rhode Island**.

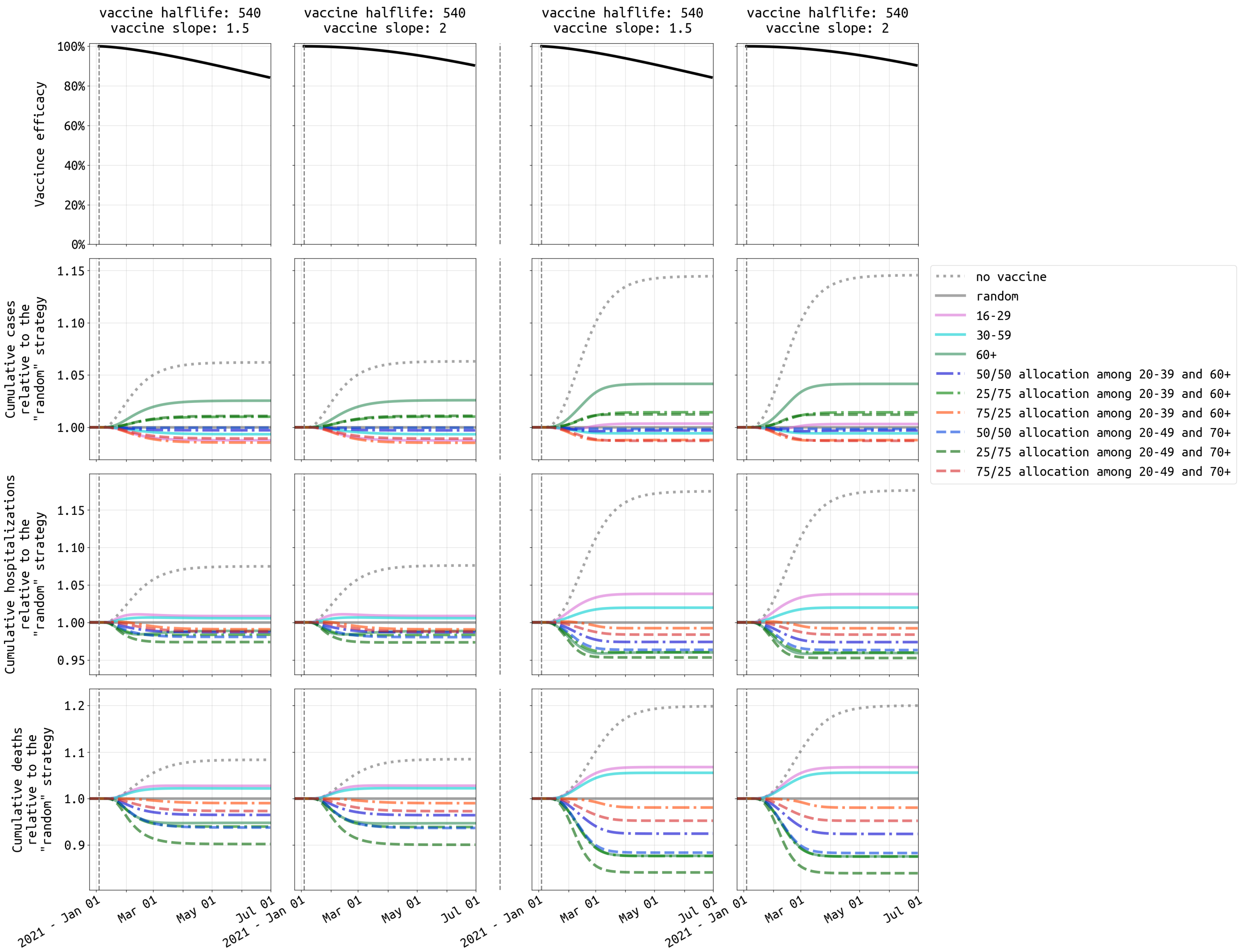


Figure S10: Similar to Figure 5, comparison of ten vaccination strategies with vaccine efficacy half-life of **540** days under **high** transmission setting in **Rhode Island**.

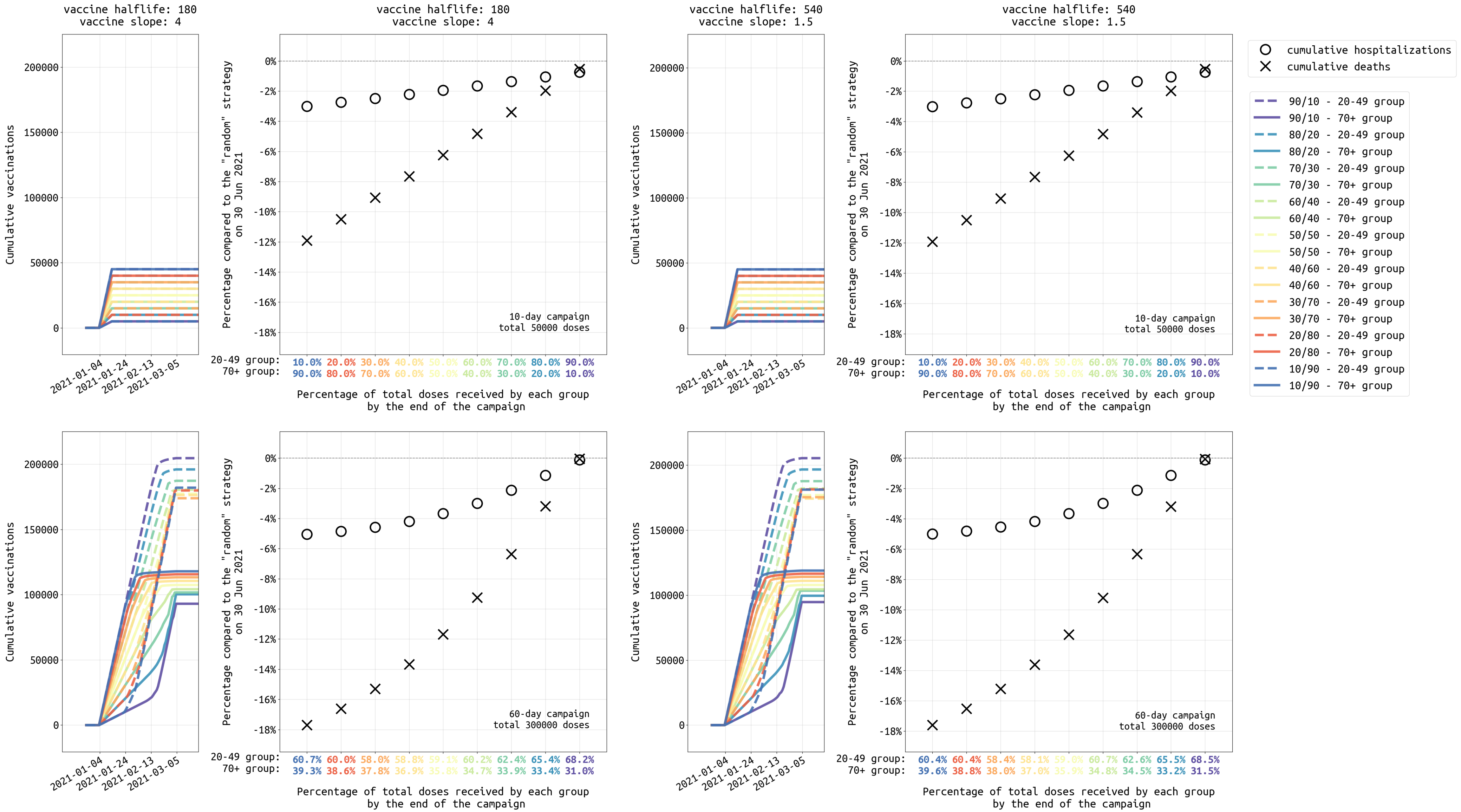


Figure S11: Similar to Figure 6, analysis of different dose allocations for strategies focused on the 20-49 and 70+ age groups, ranging from a 10/90 allocation (90% of vaccines initially given to 70+ age group) to a 90/10 allocation (90% of vaccines initially given to 20-49 age group) under **high** transmission setting in **Rhode Island**.

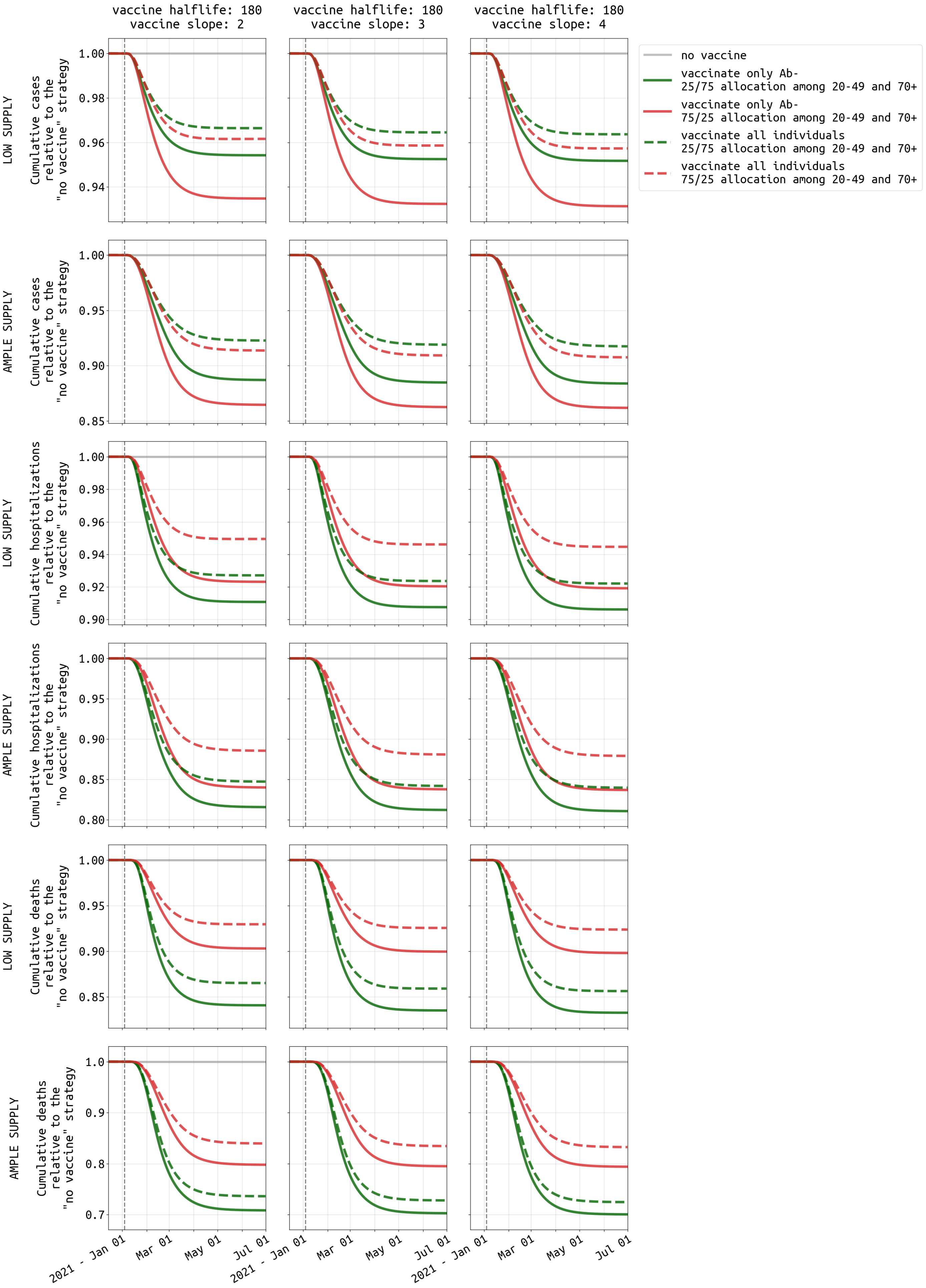


Figure S12: Similar to Figure 7, comparison of vaccinating all individuals and vaccinating only antibody-negative individuals under **high** transmission setting in **Rhode Island**.

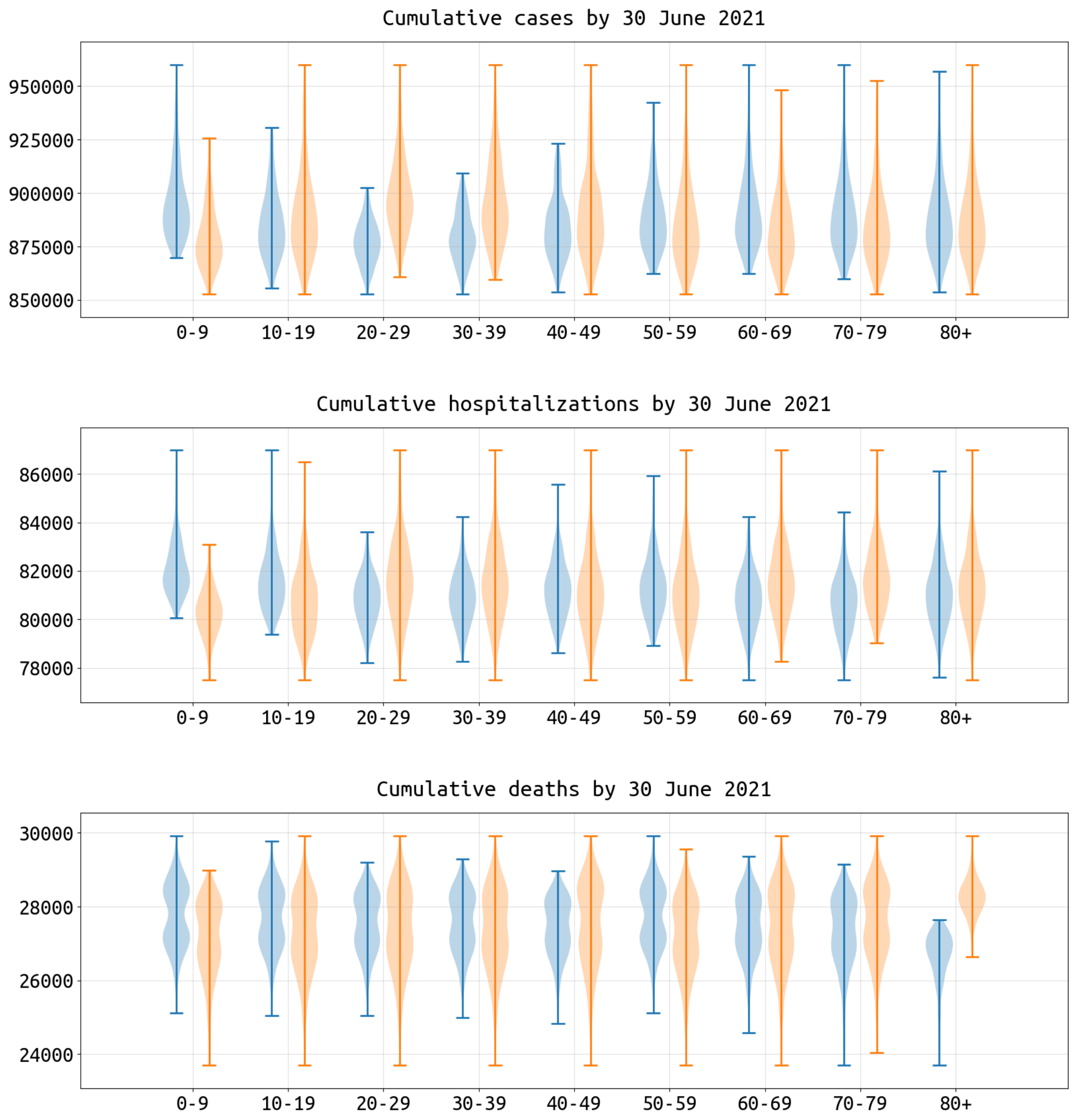


Figure S13: Similar to Figure 4, impact of including (blue) or excluding (orange) each age group in a vaccination policy, measured as reductions in cumulative cases, hospitalizations, and deaths in **Massachusetts** by 30 June 2021.

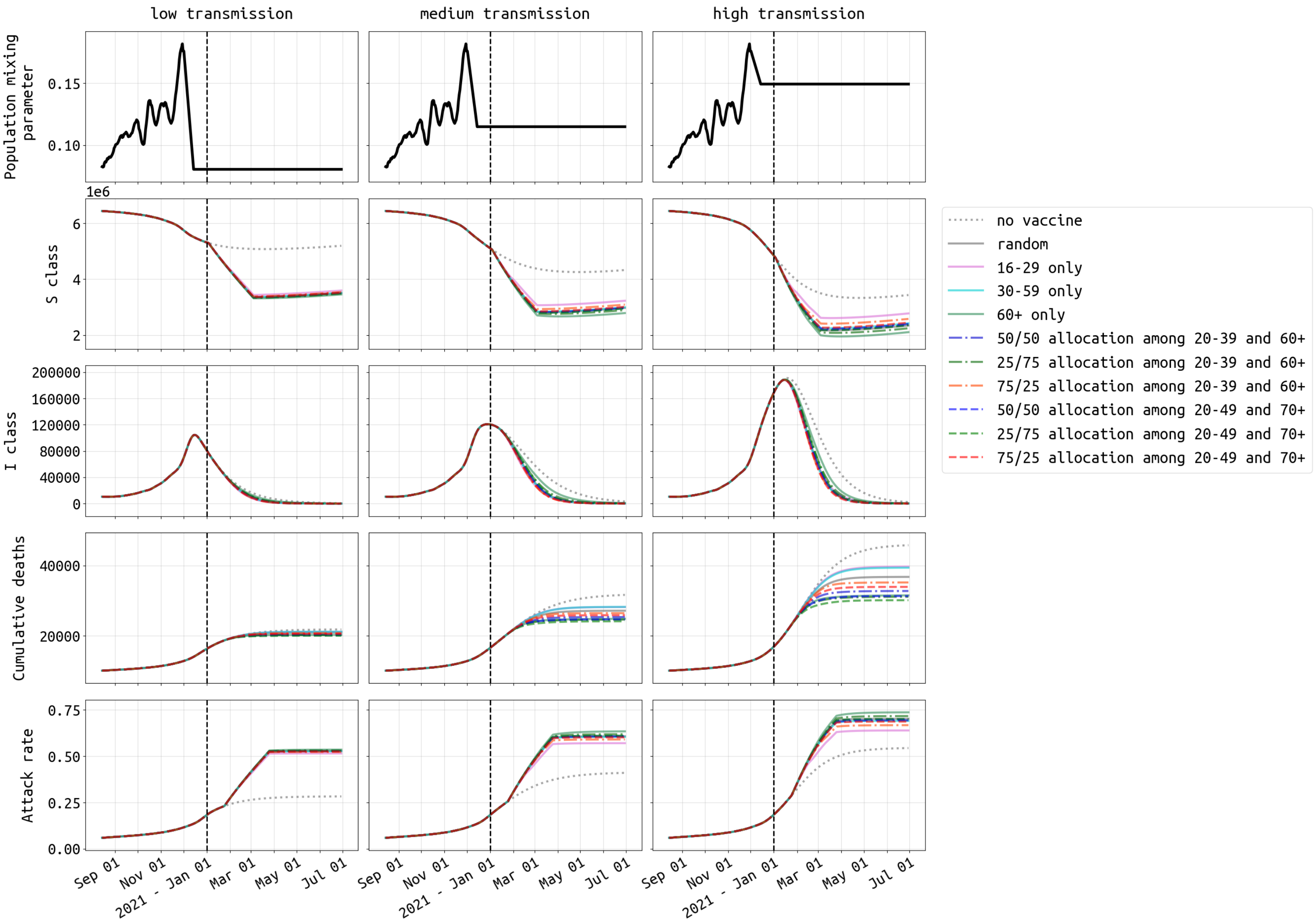


Figure S14: Simliar to Figure 3, dynamics of SARS-CoV-2 epidemic in **Massachusetts** under three different transmission scenarios and ten vaccination strategies. The vaccine profile shown here has efficacy half-life of 180 days and slope of 2. The vaccination campaign covers $1,800,000$ people (26.1% population coverage) and ends on 04 March 2021. The last row shows seroprevalence in Massachusetts from August 15, 2020 to 30 June, 2021. With no vaccination, seroprevalence would reach 28.3%, 40.1%, and 54.4% by June 30, 2021 under low, medium, and high transmission settings, respectively.

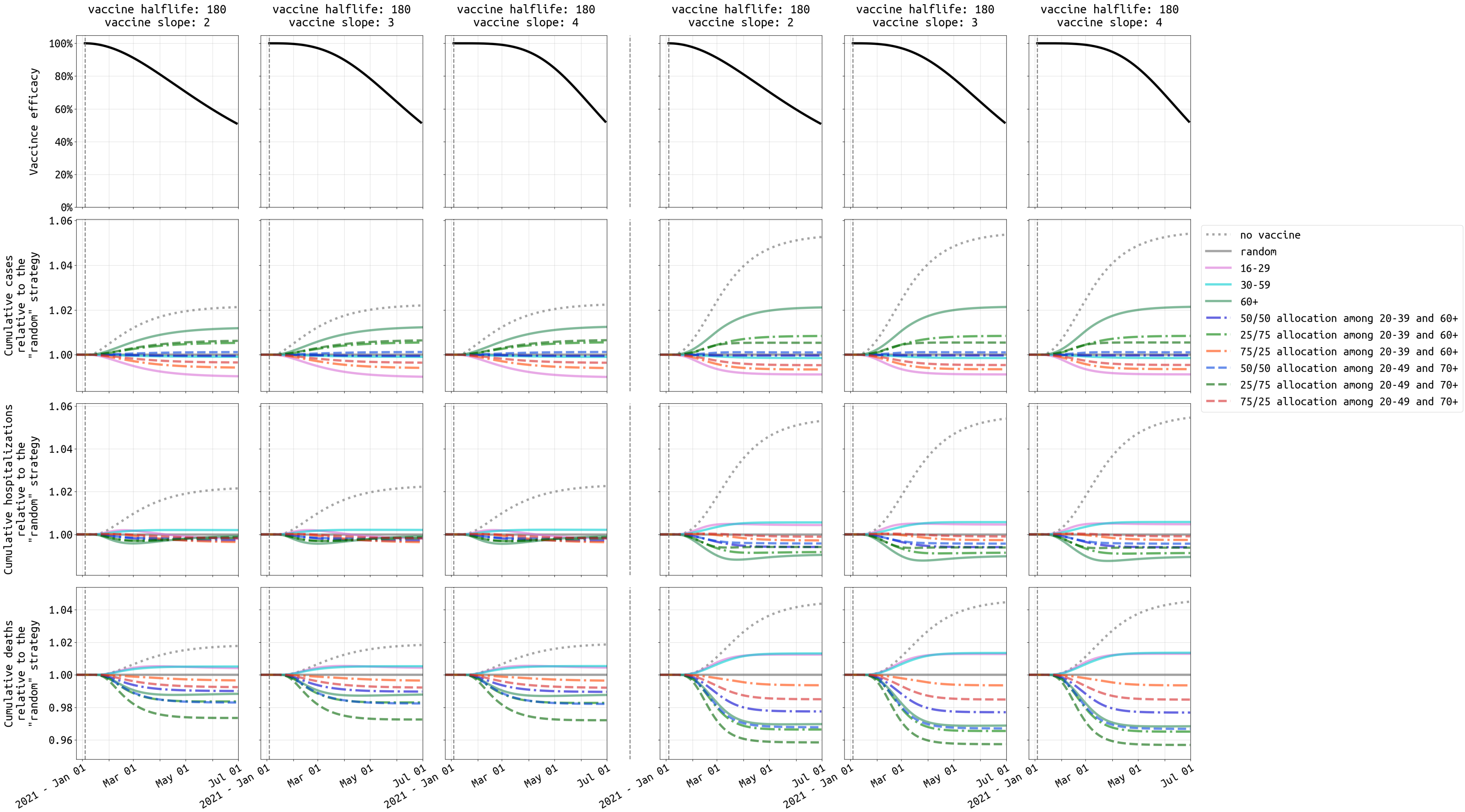


Figure S15: Similar to Figure 5, comparison of ten vaccination strategies with vaccine efficacy half-life of **180** days under **low** transmission setting in **Massachusetts**. The vaccine supply shown in the first three columns is 300,000 which is enough to cover 4.3% of Massachusetts population. The last three columns show results with 1,800,000 cumulative vaccinations (26.1% population). We use "random" (solid gray), where everybody is equally likely to receive the vaccine, as the reference strategy.

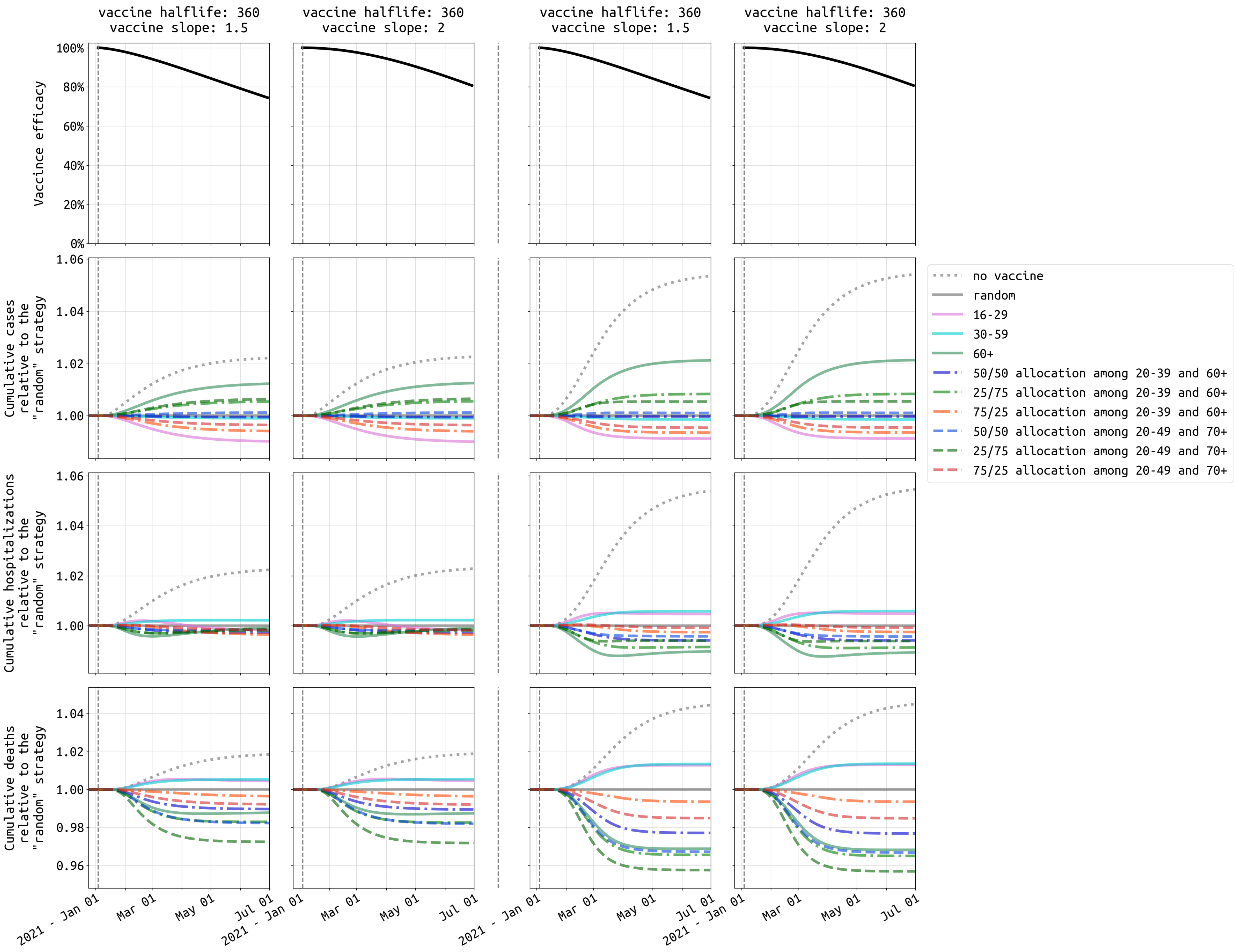


Figure S16: Similar to Figure 5, comparison of ten vaccination strategies with vaccine efficacy half-life of **360** days under **low** transmission setting in **Massachusetts**.

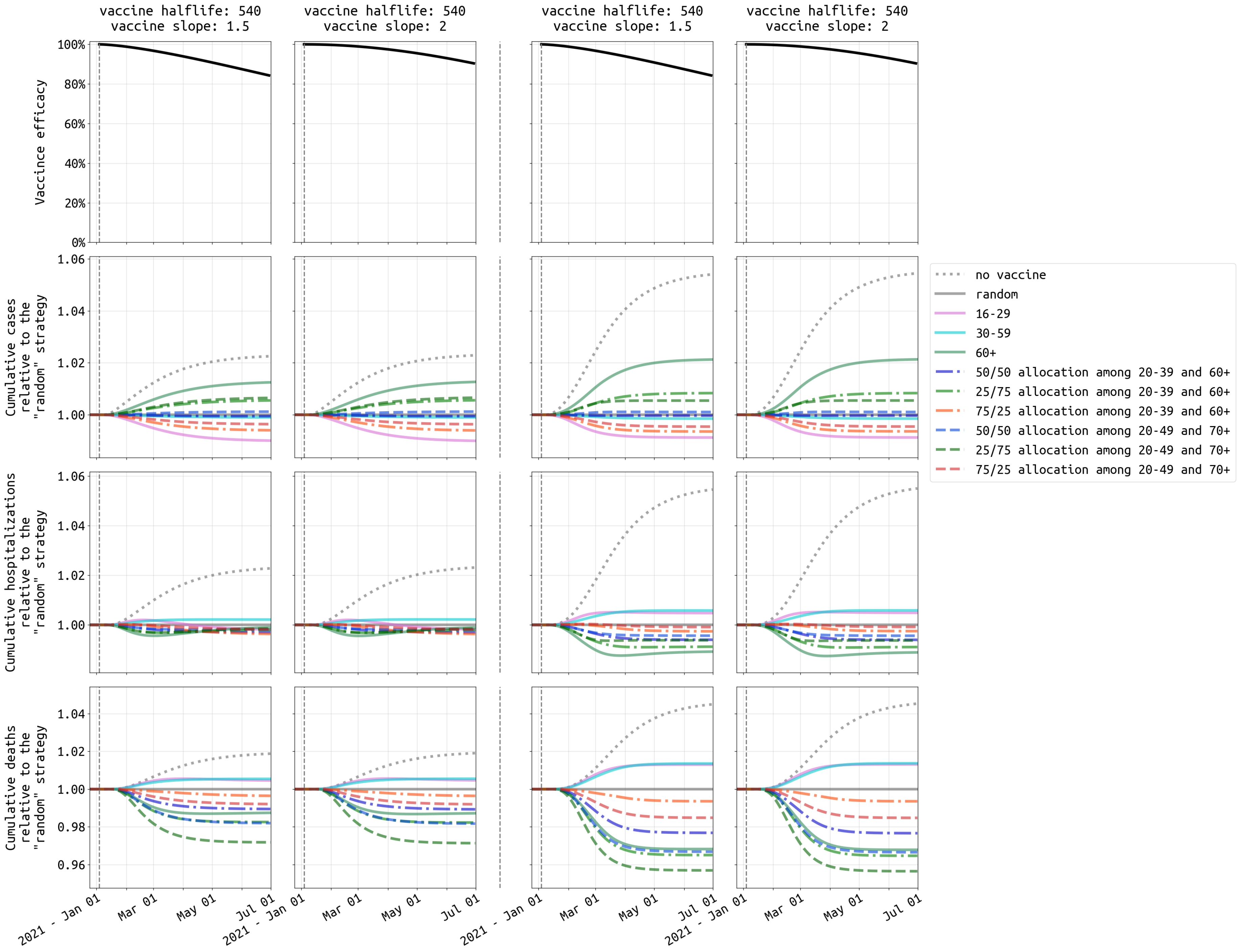


Figure S17: Similar to Figure 5, comparison of ten vaccination strategies with vaccine efficacy half-life of **540** days under **low** transmission setting in **Massachusetts**.

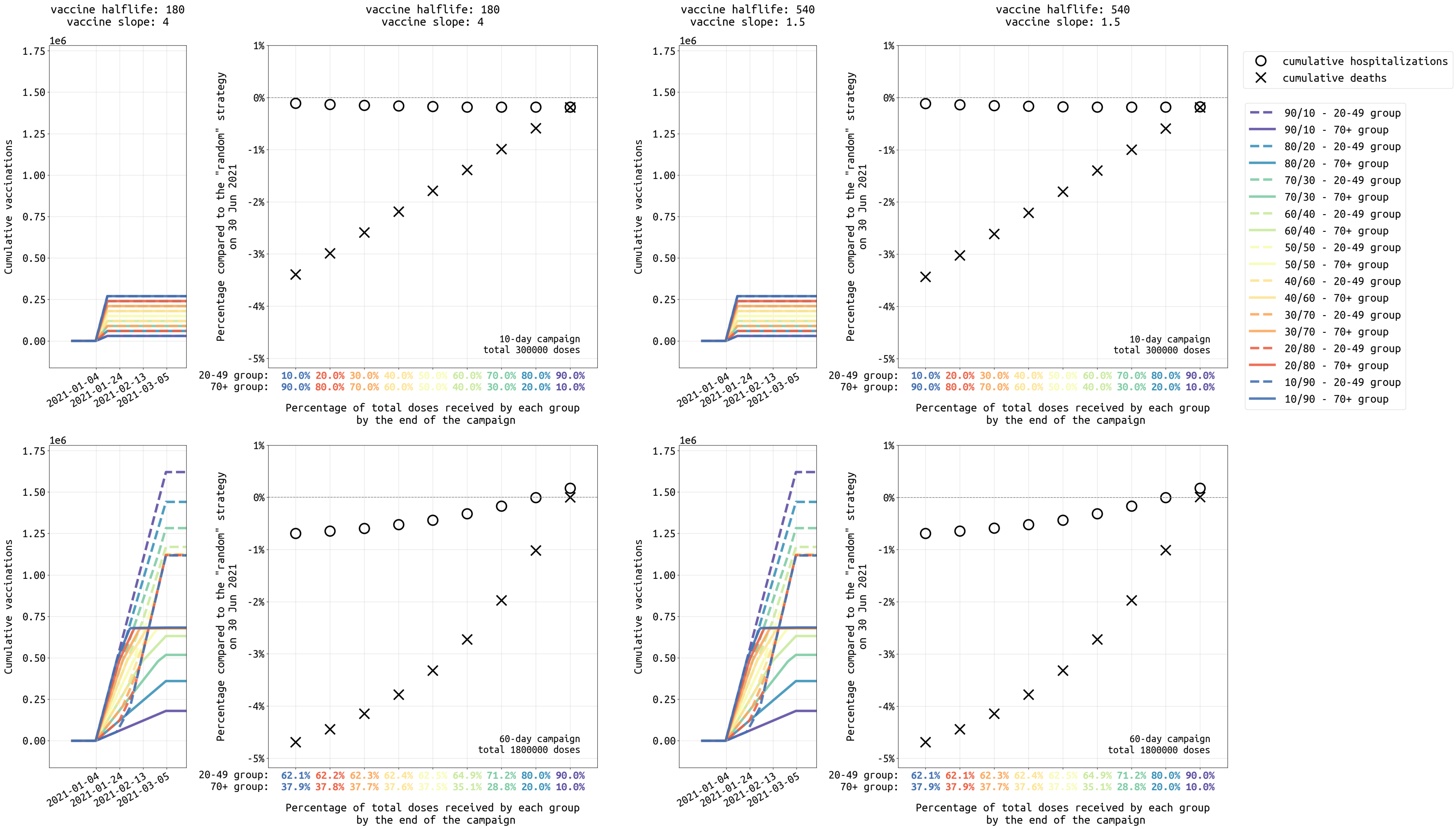


Figure S18: Similar to Figure 6, analysis of different dose allocations for strategies focused on the 20-49 and 70+ age groups, ranging from a 10/90 allocation (90% of vaccines initially given to 70+ age group) to a 90/10 allocation (90% of vaccines initially given to 20-49 age group) under **low** transmission setting in **Massachusetts**.

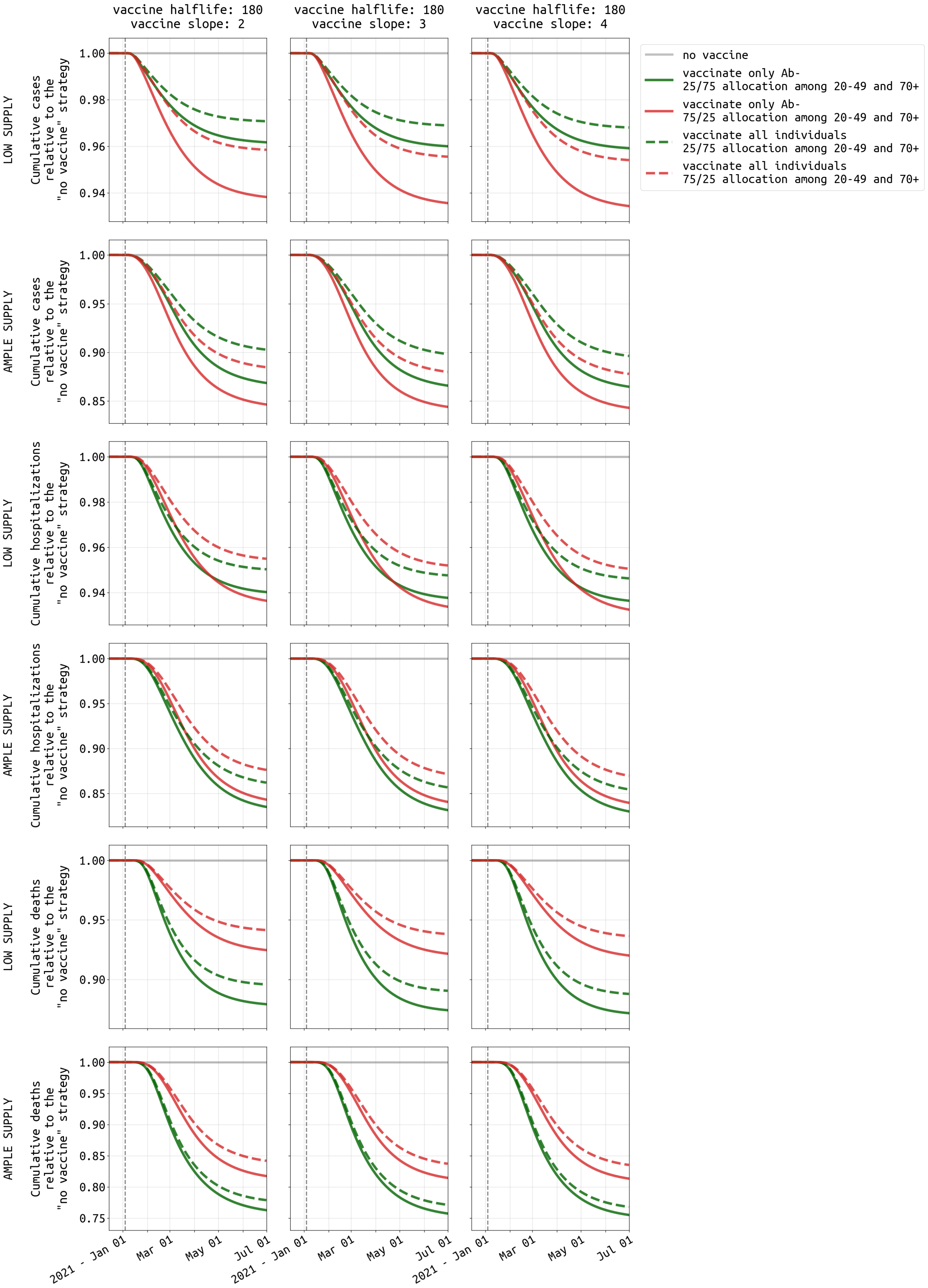


Figure S19: Similar to Figure 7, comparison of vaccinating all individuals and vaccinating only antibody-negative individuals under **low** transmission setting in **Massachusetts**.

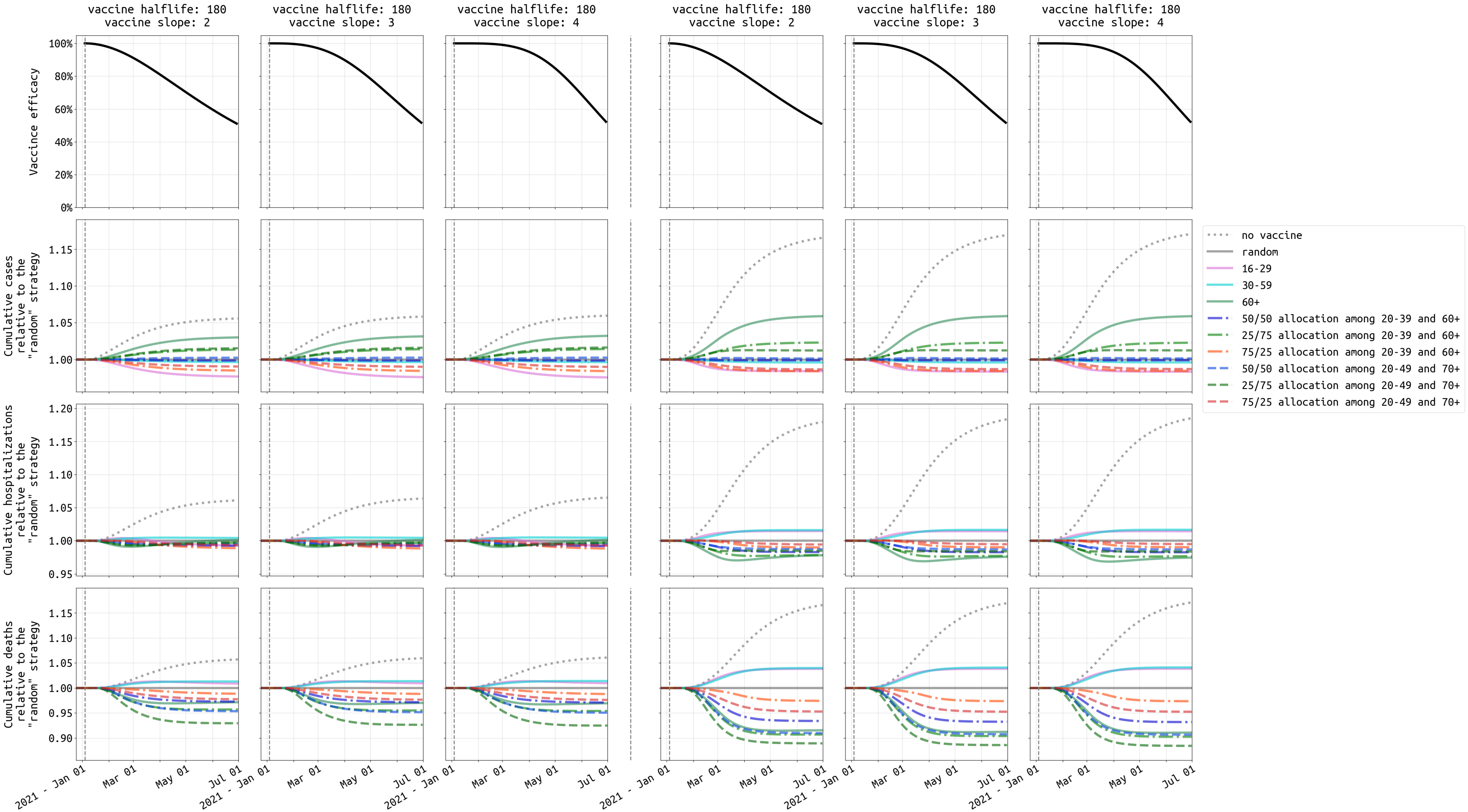


Figure S20: Similar to Figure 5, comparison of ten vaccination strategies with vaccine efficacy half-life of **180** days under **medium** transmission setting in **Massachusetts**. The vaccine supply shown in the first three columns is 300, 000 which is enough to cover 4.3% of Massachusetts population. The last three columns show results with 1, 800, 000 cumulative vaccinations (26.1% population). We use "random" (solid gray), where everybody is equally likely to receive the vaccine, as the reference strategy.

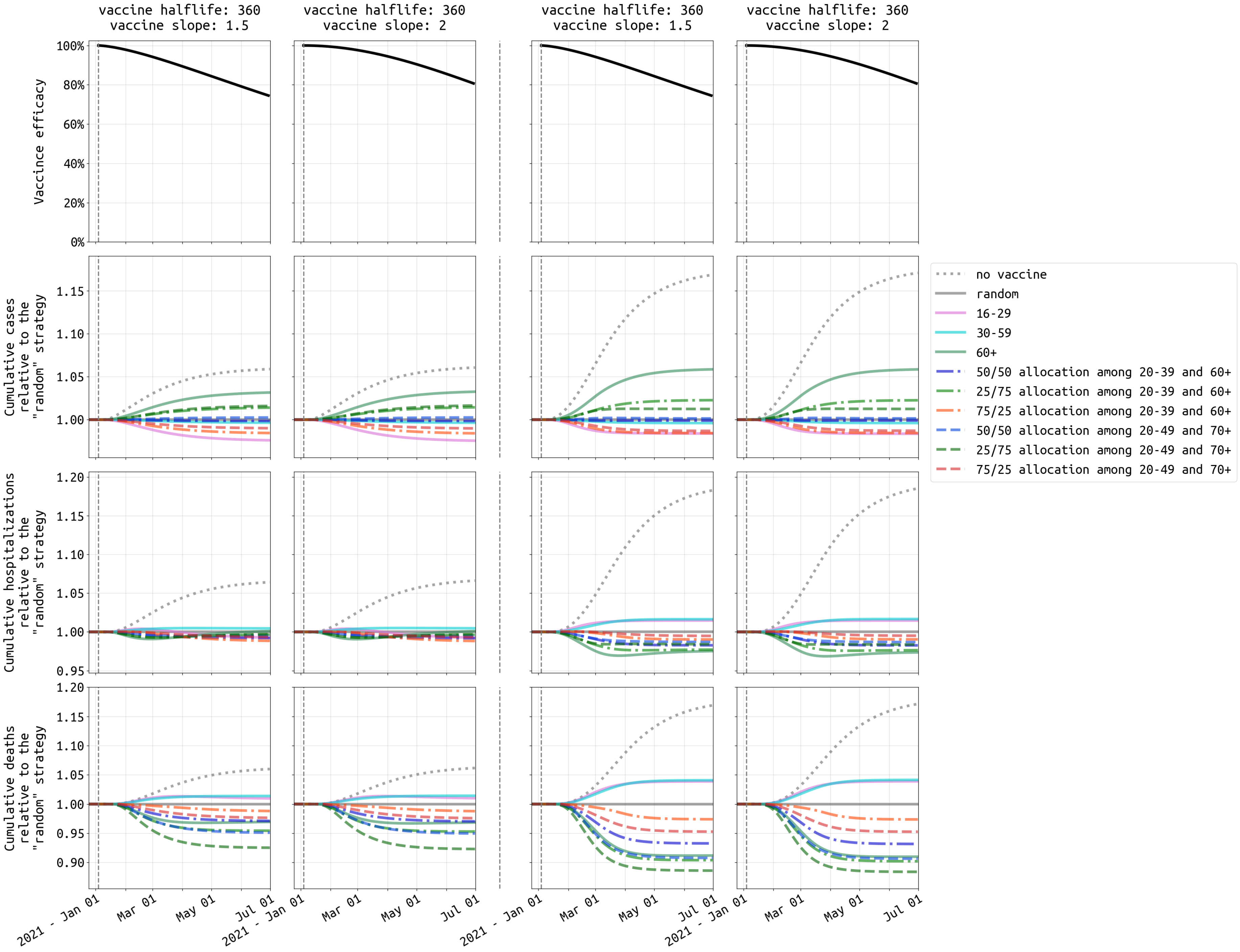


Figure S21: Similar to Figure 5, comparison of ten vaccination strategies with vaccine efficacy half-life of **360** days under **medium** transmission setting in **Massachusetts**.

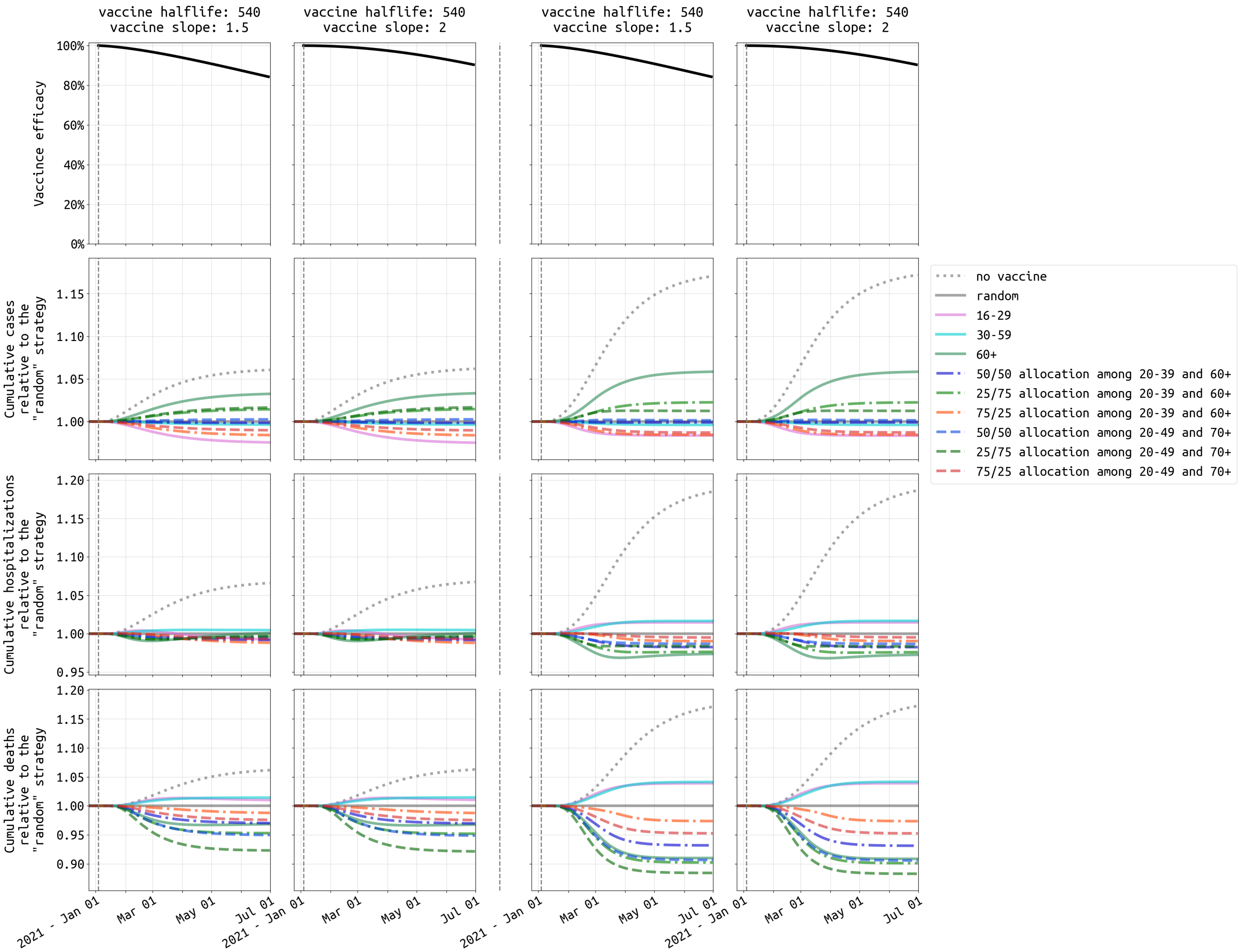


Figure S22: Similar to Figure 5, comparison of ten vaccination strategies with vaccine efficacy half-life of **540** days under **medium** transmission setting in **Massachusetts**.

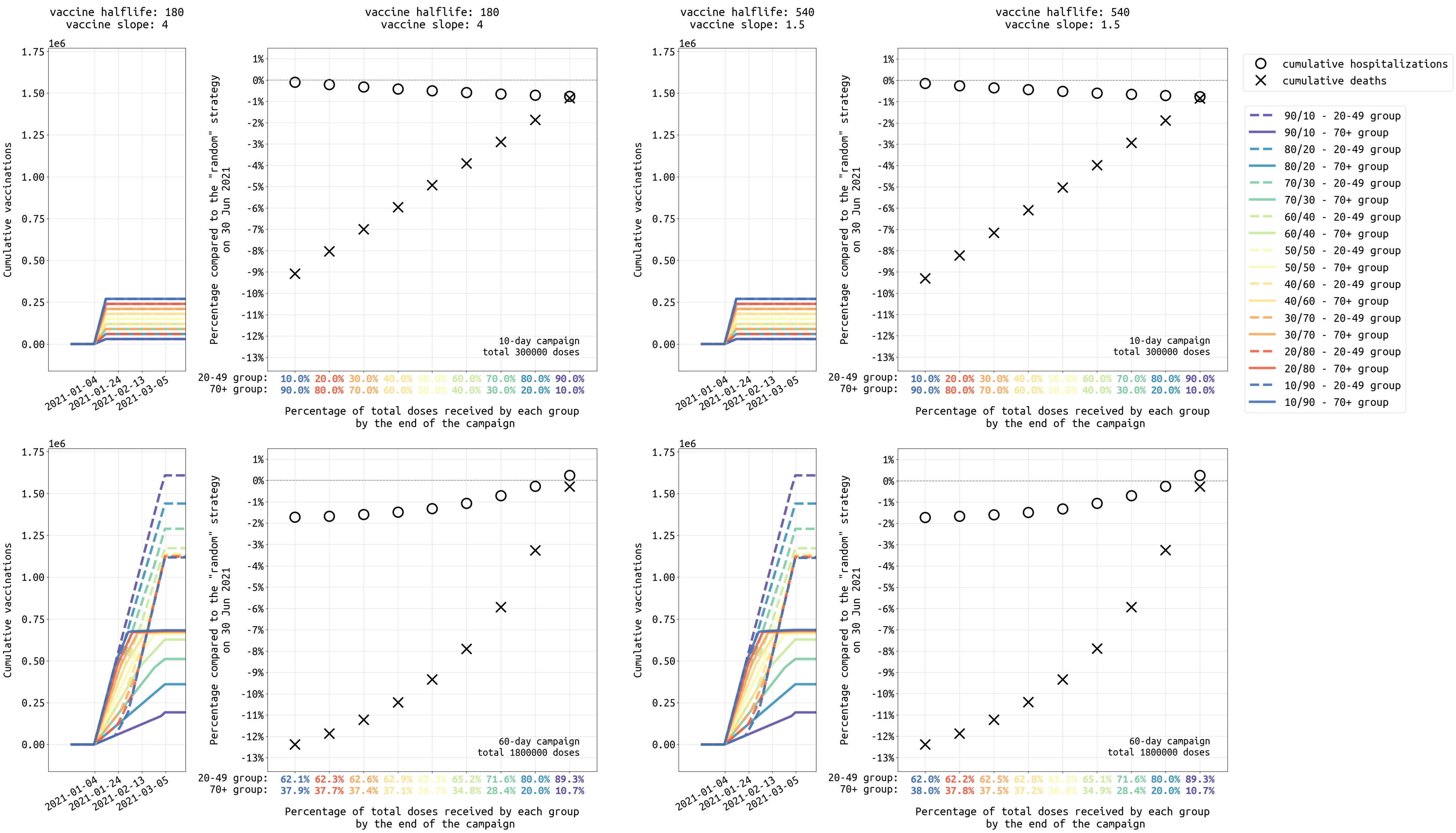


Figure S23: Similar to Figure 6, analysis of different dose allocations for strategies focused on the 20-49 and 70+ age groups, ranging from a 10/90 allocation (90% of vaccines initially given to 70+ age group) to a 90/10 allocation (90% of vaccines initially given to 20-49 age group) under **medium** transmission setting in **Massachusetts**.

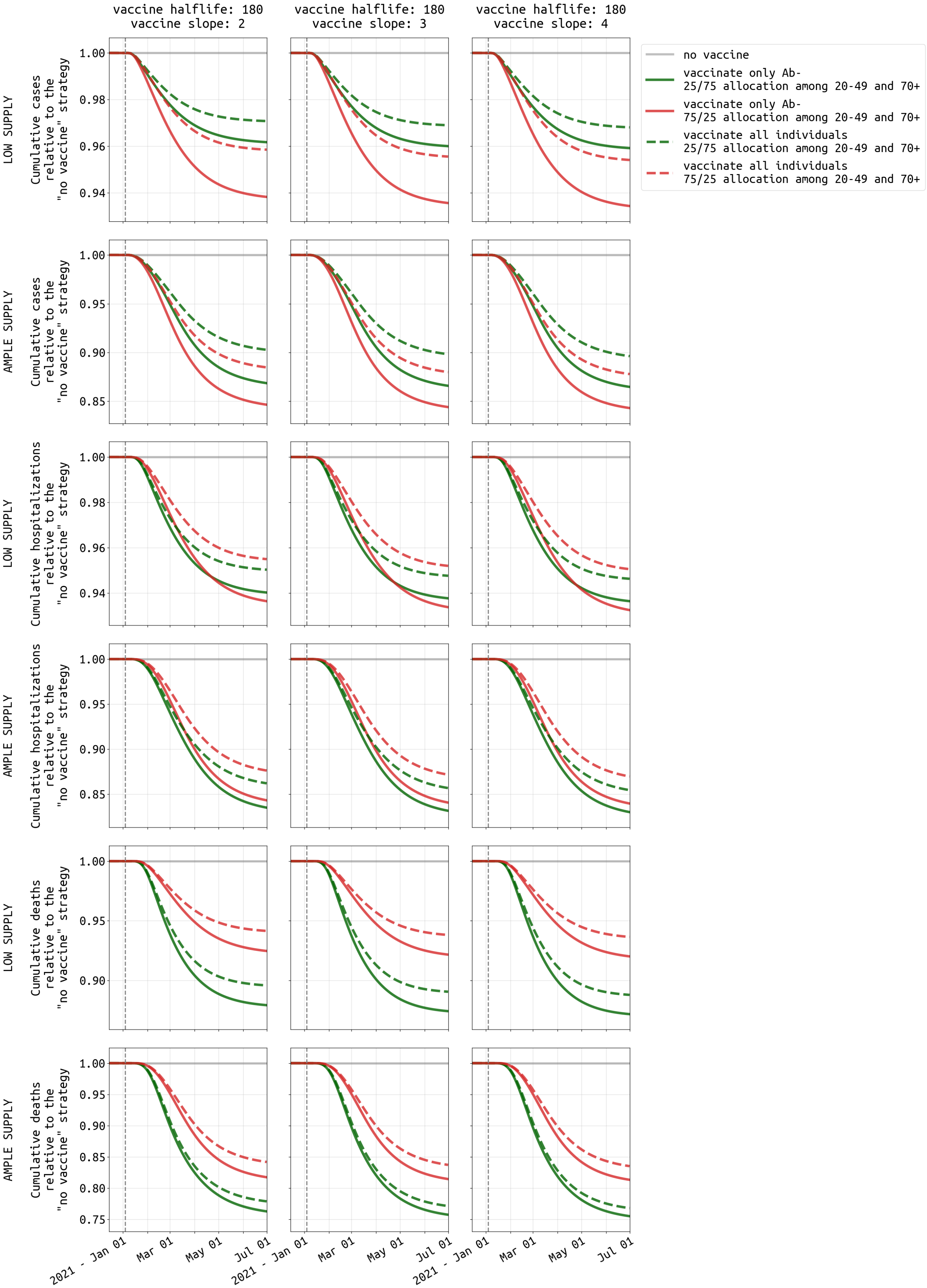


Figure S24: Similar to Figure 7, comparison of vaccinating all individuals and vaccinating only antibody-negative individuals under **medium** transmission setting in **Massachusetts**.

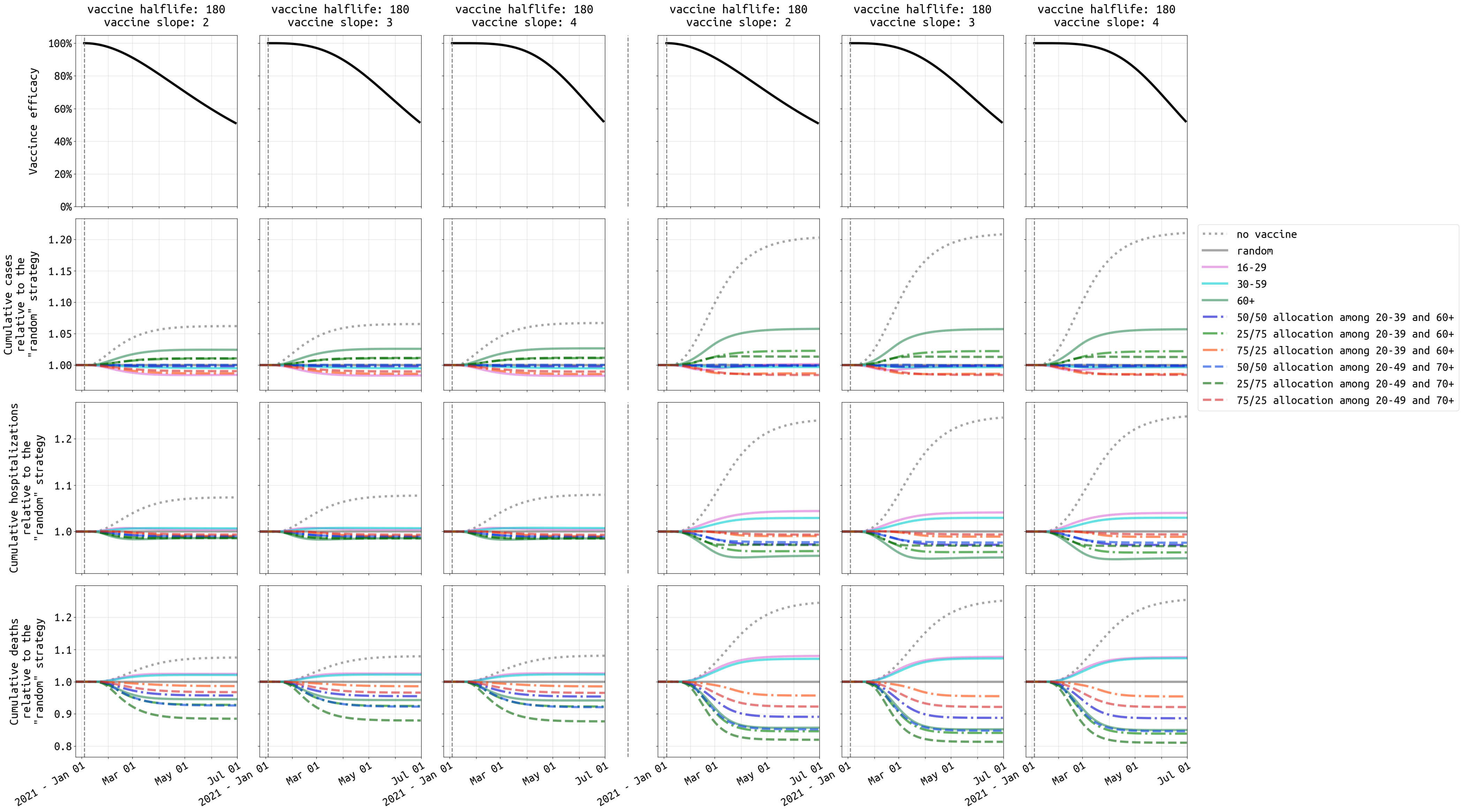


Figure S25: Similar to Figure 5, comparison of ten vaccination strategies vaccine efficacy half-life of **180** days under **high** transmission setting in **Massachusetts**. The vaccine supply shown in the first three columns is 300,000 which is enough to cover 4.3% of Massachusetts population. The last three columns show results with 1,800,000 cumulative vaccinations (26.1% population). We use "random" (solid gray), where everybody is equally likely to receive the vaccine, as the reference strategy.

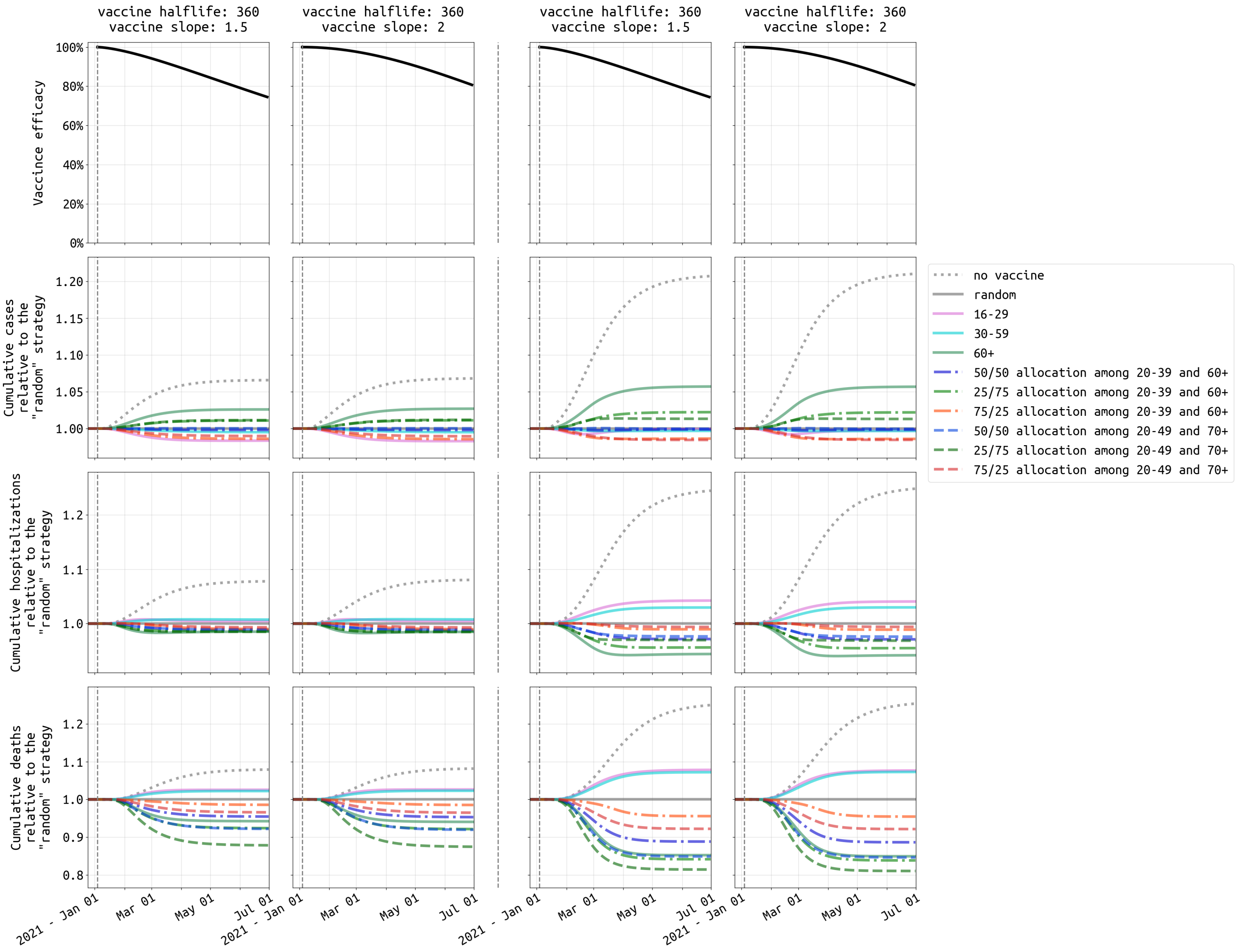


Figure S26: Similar to Figure 5, comparison of ten vaccination strategies with vaccine efficacy half-life of **360** days under **high** transmission setting in **Massachusetts**.

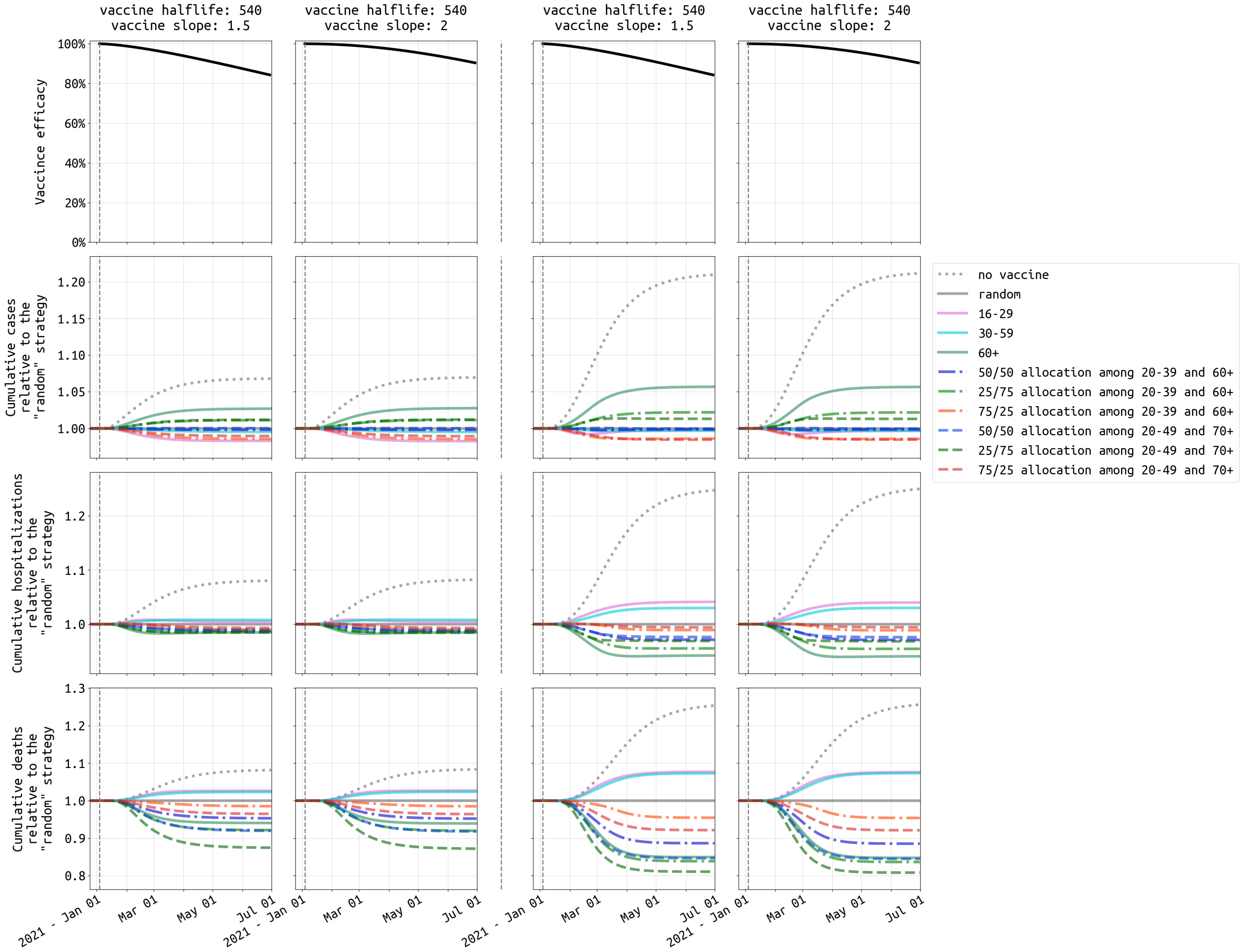


Figure S27: Similar to Figure 5, comparison of ten vaccination strategies with vaccine efficacy half-life of **540** days under **high** transmission setting in **Massachusetts**.

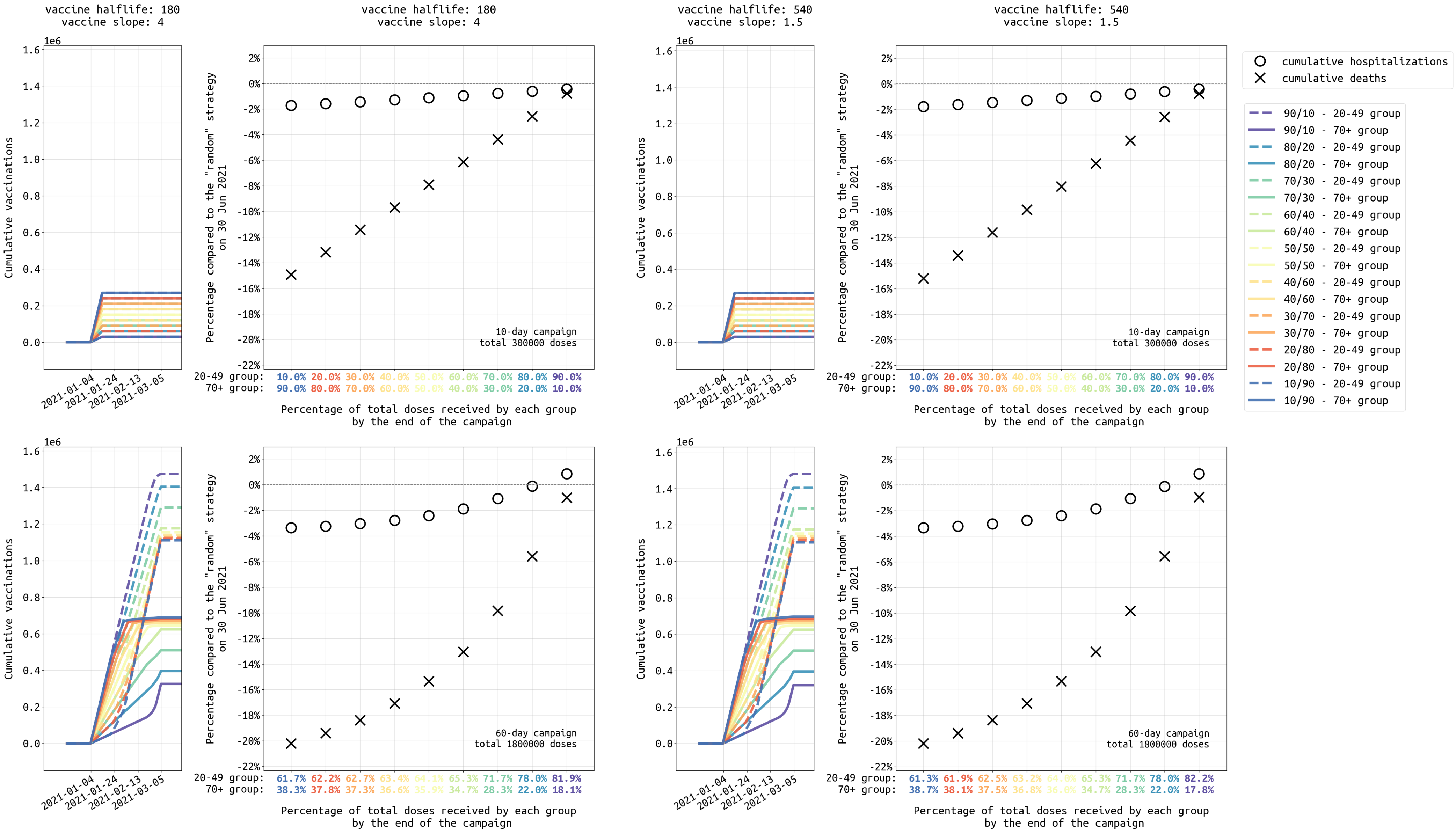


Figure S28: Similar to Figure 6, analysis of different dose allocations for strategies focused on the 20-49 and 70+ age groups, ranging from a 10/90 allocation (90% of vaccines initially given to 70+ age group) to a 90/10 allocation (90% of vaccines initially given to 20-49 age group) under **high** transmission setting in **Massachusetts**.

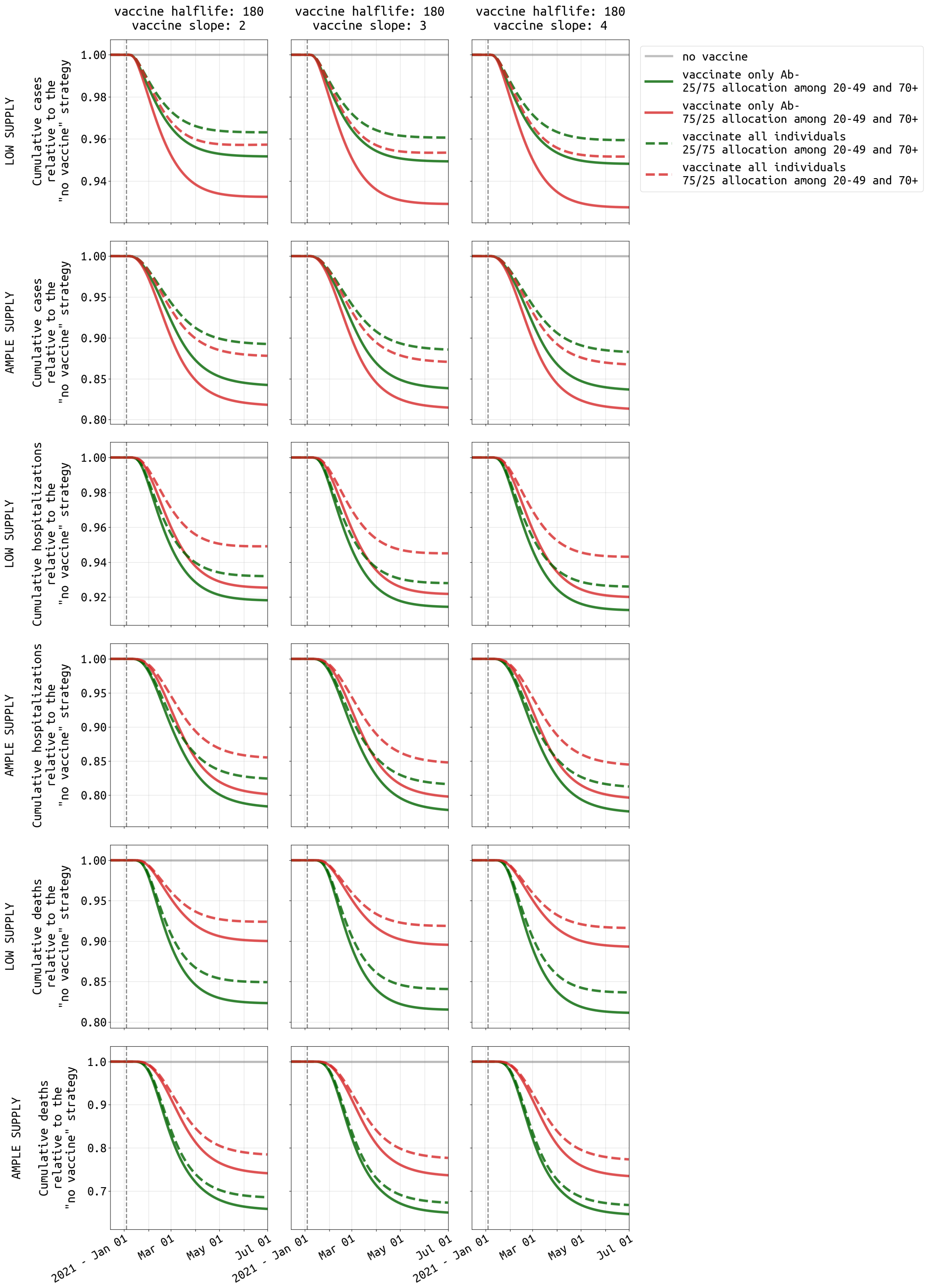


Figure S29: Similar to Figure 7, comparison of vaccinating all individuals and vaccinating only antibody-negative individuals under **high** transmission setting in **Massachusetts**.
